# COVID-19 transmission risk factors

**DOI:** 10.1101/2020.05.08.20095083

**Authors:** Alessio Notari, Giorgio Torrieri

**Author notes:** Electronic address (Alessio Notari). Electronic address (Giorgio Torrieri).

## Abstract

We analyze risk factors correlated with the initial transmission growth rate of the recent COVID-19 pandemic in different countries. The number of cases follows in its early stages an almost exponential expansion; we chose as a starting point in each country the first day *d*_*i*_ with 30 cases and we fitted for 12 days, capturing thus the early exponential growth. We looked then for linear correlations of the exponents *α* with other variables, for a sample of 126 countries. We find a positive correlation, *i.e. faster spread of COVID-19*, with high confidence level with the following variables, with respective *p*-value: low Temperature (4 · 10^−7^), high ratio of old vs. working-age people (3 · 10^−6^), life expectancy (8 · 10^−6^), number of international tourists (1· 10^−5^), earlier epidemic starting date *d*_*i*_ (2· 10^−5^), high level of physical contact in greeting habits (6 · 10^−5^), lung cancer prevalence (6 · 10^−5^), obesity in males (1· 10^−4^), share of population in urban areas (2· 10^−4^), cancer prevalence (3· 10^−4^), alcohol consumption (0.0019), daily smoking prevalence (0.0036), UV index (0.004, 73 countries). We also find a correlation with low Vitamin D serum levels (0.002*—* 0.006), but on a smaller sample, ~ 50 countries, to be confirmed on a larger sample. There is highly significant correlation also with blood types: positive correlation with types RH-(3· 10^−5^) and A+ (3 ·10^−3^), negative correlation with B+ (2 ·10^−4^). We also find positive correlation with moderate confidence level (*p*-value of 0.02∼ 0.03) with: CO_2_/SO emissions, type-1 diabetes in children, low vaccination coverage for Tuberculosis (BCG). Several of the above variables are correlated with each other and so they are likely to have common interpretations. We thus performed a Principal Component Analysis, in order to find the significant independent linear combinations of such variables. We also analyzed the possible existence of a bias: countries with low GDP-per capita might have less intense testing and we discuss correlation with the above variables.

## I. INTRODUCTION

The recent coronavirus (COVID-19) pandemic is now spreading essentially everywhere in our planet. The growth rate of the contagion has however a very high variability among different countries, even in its very early stages, when government intervention is still almost negligible. Any factor contributing to a faster or slower spread needs to be identified and understood with the highest degree of scrutiny. In [1] the early growth rate of the contagion has been found to be correlated at high significance with temperature T. In this work we extend a similar analysis to many other variables. This correlational study could help further investigation in order to establish causal factors and it can help policy makers in their decisions.

Some factors are intuitive and have been found in other studies, such as temperature [1, 9] (see also [10] for a different conclusion) and air travel [2, 10]; we aim here at being more exhaustive and at finding also factors which are not “obvious” and have a potential biological origin, or correlation with one.

The paper is organized as follows. In section II we explain our methods, in section III we show our main results, in section IV we show the detailed results for each individual variable of our analysis, in section V we discuss correlations among variables and in section VI we draw our conclusions.

## II. METHOD

As in [1], we use the empirical observation that the number of COVID-19 positive cases follows a common pattern in the majority of countries: once the number of confirmed cases reaches order 10 there is a very rapid growth, which is typically well approximated for a few weeks by an exponential. Subsequently the exponential growth typically gradually slows down, probably due to other effects, such as: lockdown policies from governments, a higher degree of awareness in the population or the tracking and isolation of the positive cases. The growth is then typically stopped and reaches a peak in countries with a strong lockdown/tracking policy.

Our aim is to find which factors correlate with the speed of contagion, in its first stage of *free* propagation. For this purpose we analyzed a datasets of 126 countries taken from [12] on April 15th. We have chosen our sample using the following rules:

- We start analyzing data from the first day *d*_*i*_ in which the number of cases in a given country reaches a reference number *N*_*i*_, which we choose to be *N*_*i*_ = 30 [57];
- We include only countries with at least 12 days of data, after this starting point;
- We excluded countries with too small total population (less than 300 thousands inhabitants).

We then fit the data for each country with a simple exponential curve *N* (*t*) = *N*_0_ *e*^*αt*^, with 2 parameters, *N*_0_ and *α*; here *t* is in units of days.

Note that the statistical errors on the exponents *α* are typically only a few percent of the spread of the values of *α* among the various countries. For this reason we disregarded statistical errors on *α*. The analysis was done using the software *Mathematica*, from Wolfram Research, Inc..

## III. MAIN RESULTS

We first look for correlations with several individual variables. Most variables are taken from [13], while for a few of them have been collected from other sources, as commented below.

### 1. Non-significant variables

We find *no* significant correlation of the COVID-19 transmission in our set of countries with many variables, including the following ones:

1. Number of inhabitants;
2. Asthma-prevalence;
3. Participation time in leisure, social and associative life per day;
4. Population density;
5. Average precipitation per year;
6. Vaccinations coverage for: Polio, Diphteria, Tetanus, Pertussis, Hepatitis B;
7. Share of men with high-blood-pressure;
8. Diabetes prevalence (type 1 and 2, together);
9. Air pollution (“Suspended particulate matter (SPM), in micrograms per cubic metre”).

### 2. Significant variables, strong evidence

We find *strong* evidence for correlation with:

1. Temperature (negative correlation, *p*-value 4.4 · 10^−7^);
2. Old-age dependency ratio: ratio of the number of people older than 64 relative to the number of people in the working-age (15-64 years) (positive correlation, *p*-value 3.3 · 10^−6^);
3. Life expectancy (positive correlation *p*-value 8.1 · 10^−6^);
4. International tourism: number of arrivals (positive correlation *p*-value 9.6 · 10^−6^);
5. Starting day *d*_*i*_ of the epidemic (negative correlation, *p*-value 1.7 · 10^−5^);
6. Amount of contact in greeting habits (positive correlation, *p*-value 5.0 · 10^−5^);
7. Lung cancer death rates (positive correlation, *p*-value 6.3 · 10^−5^);
8. Obesity in males (positive correlation, *p*-value 1.2 · 10^−4^);
9. Share of population in urban areas (positive correlation, *p*-value 1.7 · 10^−4^);
10. Share of population with cancer (positive correlation, *p*-value 2.8 · 10^−4^);
11. Alcohol consumption (positive correlation, *p*-value 0.0019);
12. Daily smoking prevalence (positive correlation, *p*-value 0.0036);
13. UV index (negative correlation, *p*-value 0.004; smaller sample, 73 countries);
14. Vitamin D serum levels (negative correlation, annual values *p*-value 0.006, seasonal values 0.002; smaller sample, ∼ 50 countries).

### 3. Blood types, strong evidence

We also find strong evidence for correlation with blood types:

1. RH + blood group system (negative correlation, *p*-value 3 · 10^−5^);
2. A+ (positive correlation, *p*-value 3 · 10^−3^);
3. B + (negative correlation, *p*-value 2 · 10^−4^);
4. A-(positive correlation, *p*-value 3 · 10^−5^);
5. 0-(positive correlation, *p*-value 8 · 10^−4^);
6. AB-(positive correlation, *p*-value 0.028).

We find moderate evidence for correlation with:

1. B-(positive correlation, *p*-value 0.013).

We find no significant correlation with:

1. 0+;
2. AB+.

### 4. Significant variables, moderate evidence

We find moderate evidence for correlation with:

1. CO_2_ (and SO) emissions (positive correlation, *p*-value 0.015);

2. Type-1 diabetes in children (positive correlation, *p*-value 0.023);

3. Vaccination coverage for Tuberculosis (BCG) (negative correlation, *p*-value 0.028).

### 5. Significant variables, counterintuitive

Counterintuitively we also find correlations in a direction opposite to a naive expectation:

1. Death-rate-from-air-pollution (negative correlation, *p*-value 3.5 · 10^−5^);
2. Prevalence of anemia, adults and children, (negative correlation, *p*-value 1.4 ·10^−4^ and 7.*×*10^−6^, respectively);
3. Share of women with high-blood-pressure (negative correlation, *p*-value 1.6 · 10^−4^);
4. Incidence of Hepatitis B (negative correlation, *p*-value 2.4 · 10^−4^);
5. PM2.5 air pollution (negative correlation, *p*-value 0.029).

#### A. Bias due to GDP: lack of testing?

We also find a correlation with GDP per capita, which should be an indicator of lack of testing capabilities. Note however that GDP per capita is also quite highly correlated with another important variable, life expectancy, as we will show in section V: high GDP per capita is related to an older population, which is correlated with faster contagion.

Note also that correlation of contagion with GDP disappears when excluding very poor countries, approximately below 5 thousand $ GDP per capita: this is likely due to the fact that only below a given threshold the capability of testing becomes insufficient.

We performed 2-variables fits, including GDP and each of the above significant variables, in order to check if they remain still significant. In section IV we will show the results of such fits, and also the result of individual one variable fits excluding countries below the threshold of 5 thousand $ GDP per capita. We list here below the variables that are still significant even when fitting together with GDP.

### 1. Significant variables, strong evidence

In a 2-variable fit, including GDP per capita, we find strong evidence for correlation with:

1. Amount of contact in greeting habits (positive correlation, *p*-value 1.5 · 10^−5^);
2. Temperature (negative correlation, *p*-value 2.3 · 10^−5^);
3. International tourism: number of arrivals (positive correlation, *p*-value 2.6 · 10^−4^);
4. Old-age dependency ratio: ratio of the number of people older than 64 relative to the number of people in the working-age (15-64 years) (positive correlation, *p*-value 5.5 · 10^−4^);

5. Vitamin D serum levels (negative correlation, annual values *p*-value 0.0032, seasonal values 0.0024; smaller sample, ∼ 50 countries). To be confirmed on a larger sample.

6. Starting day of the epidemic (negative correlation, *p*-value 0.0037);

7. Lung cancer death rates (positive correlation, *p*-value 0.0039);

8. Life expectancy (positive correlation, *p*-value 0.0048);

### 2. Blood types, strong evidence

We still find strong evidence for correlation with blood types:

1. RH + blood group system (negative correlation, *p*-value 1 · 10^−3^);
2. B + (negative correlation, *p*-value 2 · 10^−3^);
3. A-(positive correlation, *p*-value 1 · 10^−3^);
4. 0-(positive correlation, *p*-value 3 · 10^−3^);

We find moderate evidence for correlation with:

1. A+ (positive correlation, *p*-value 0.028);
2. B-(positive correlation, *p*-value 0.039).
3. AB-(positive correlation, *p*-value 0.012).

### 3. Significant variables, moderate evidence

We find moderate evidence for:

1. UV index (negative correlation, *p*-value 0.01; smaller sample, 73 countries);
2. Type-I diabetes in children, 0-19 years-old (negative correlation, *p*-value 0.01);
3. Vaccination coverage for Tuberculosis (BCG) (negative correlation, *p*-value 0.023);
4. Obesity in males (positive correlation, *p*-value 0.02);
5. CO_2_ emissions (positive correlation, *p*-value 0.02);
6. Alcohol consumption (positive correlation, *p*-value 0.03);
7. Daily smoking prevalence (positive correlation, *p*-value 0.03);
8. Share of population in urban areas (positive correlation, *p*-value 0.04);

### 4. Significant variables, counterintuitive

Counterintuitively we still find correlations with:

1. Death rate from air pollution (negative correlation, *p*-value 0.002);
2. Prevalence of anemia, adults and children, (negative correlation, *p*-value 0.023 and 0.005);
3. Incidence of Hepatitis B (negative correlation, *p*-value 0.01);
4. Share of women with high-blood-pressure (negative correlation, *p*-value 0.03).

In the next section we analyze in more detail the significant variables, one by one (except for those which are not significant anymore after taking into account of GDP per capita). In section V we will analyze cross-correlations among such variables and this will also give a plausible interpretation for the existence of the “counterintuitive” variables.

## IV. RESULTS FOR EACH VARIABLE

We first analyze each individual variable, then we analyze variables that have a “counterintuitive” correlation and finally we separately analyze blood types.

### 1. Temperature

The average temperature T has been collected for the relevant period of time, ranging from January to mid April, weighted among the main cities of a given country, see [1] for details. Results are shown in fig. 2 and Table II.

**Table I:** In the left panel: best-estimate, standard error (*σ*), t-statistic and *p*-value for the parameters of the linear interpolation, for correlation of *α* with GDP per capita (*GDP*). In the right panel: *R*^2^ for the best-estimate and number of countries *N*.

| | Estimate | Standard Error | t-Statistic | $p$-value | | |
| --- | --- | --- | --- | --- | --- | --- |
| 1 | 0.155 | 0.0111 | 14. | $6.79 \times 10^{-27}$ | $R^2$ | 0.087 |
| $GDP$ | $1.28 \times 10^{-6}$ | $3.81 \times 10^{-7}$ | 3.37 | 0.001 | $N$ | 121 |

**Table II:**
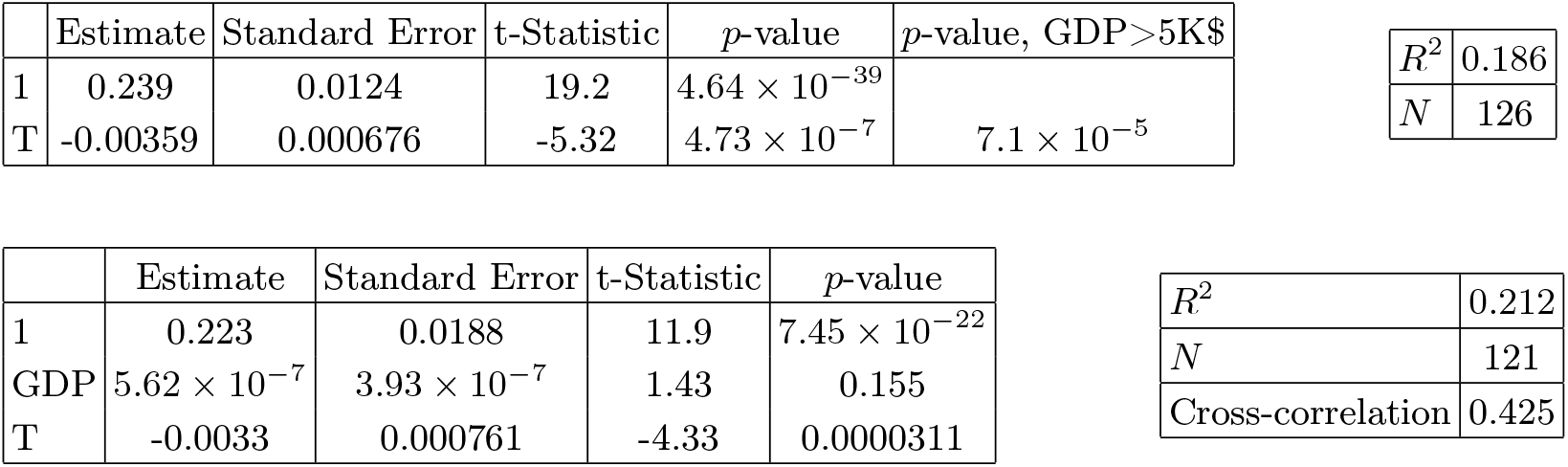
In the left top panel: best-estimate, standard error (*σ*), t-statistic and *p*-value for the parameters of the linear interpolation, for correlation of *α* with temperature T. We also show the *p*-value, excluding countries below 5 thousand $ GDP per capita. In the left bottom panel: same quantities for correlation of *α* with temperature T and GDP per capita. In the right panels: *R*^2^ for the best-estimate and number of countries *N*. We also show the correlation coefficient between the 2 variables in the two-variable fit.

**Figure 1:**
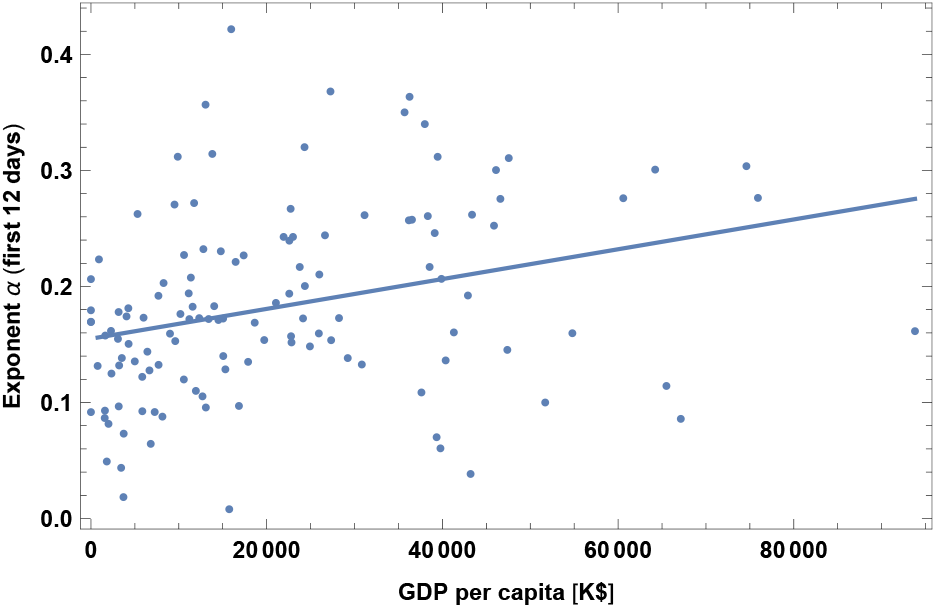
Exponent *α* for each country vs. GDP per capita. We show the data points and the best-fit for the linear interpolation.

**Figure 2:**
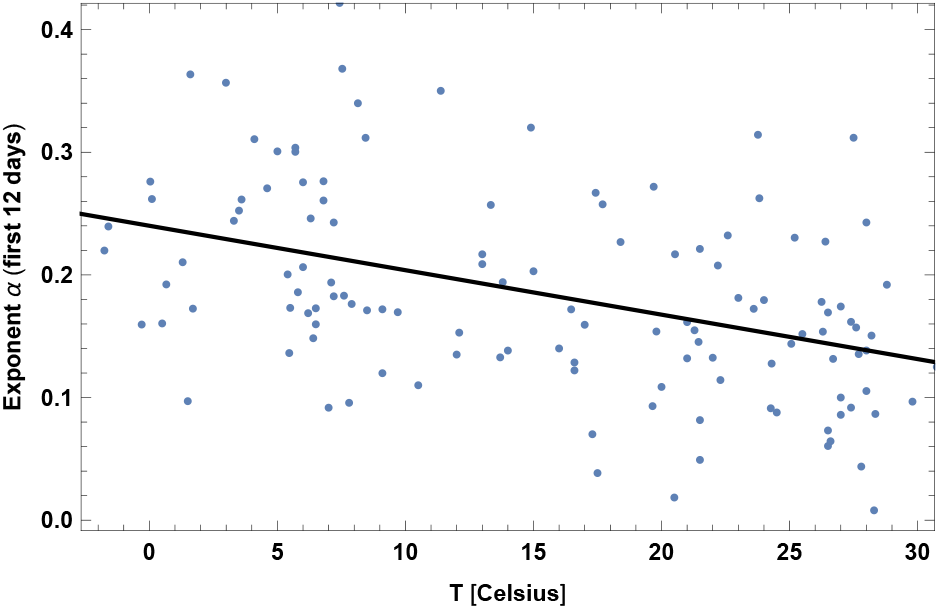
Exponent *α* for each country vs. average temperature T, for the relevant period of time, as defined in [1]. We show the data points and the best-fit for the linear interpolation.

We also found that another variable, the absolute value of latitude, has a similar amount of correlation as for the case of T. However the two variables have a very high correlation (about 0.91) and we do not show results for latitude here. Another variable which is also very highly correlated is UV index and it is shown later.

### 2. Old-age dependency ratio

This is the ratio of the number of people older than 64 relative to the number of people in the working-age (15-64 years). Data are shown as the proportion of dependents per 100 working-age population, for the year 2017. Results are shown in fig. 3 and Table III. This is an interesting finding, since it may suggest that old people are not only subject to higher mortality, but also more likely to be contagious. This could be either because they are more likely to become sick, or because their state of sickness is longer and more contagious, or because many of them live together in nursing homes, or all such reasons together.

**Table III:** In the left top panel: best-estimate, standard error (*σ*), t-statistic and *p*-value for the parameters of the linear interpolation, for correlation of *α* with old-age dependency ratio, OLD. We also show the *p*-value, excluding countries below 5 thousand $ GDP per capita. In the left bottom panel: same quantities for correlation of *α* with OLD and GDP per capita. In the right panels: *R*^2^ for the best-estimate and number of countries *N*. We also show the correlation coefficient between the 2 variables in the two-variable fit.

| | Estimate | Standard Error | t-Statistic | $p$-value | $p$-value, GDP>5K\$ | | |
| --- | --- | --- | --- | --- | --- | --- | --- |
| 1 | 0.132 | 0.0126 | 10.5 | $9.03 \times 10^{-19}$ | | $R^2$ | 0.164 |
| OLD | 0.00326 | 0.000669 | 4.87 | $3.37 \times 10^{-6}$ | 0.0025 | $N$ | 123 |

Table III: In the left top panel: best-estimate, standard error ( $\sigma$ ), t-statistic and $p$ -value for the parameters of the linear interpolation, for correlation of $\alpha$ with old-age dependency ratio, OLD.
| | Estimate | Standard Error | t-Statistic | $p$-value | | |
| --- | --- | --- | --- | --- | --- | --- |
| 1 | 0.126 | 0.0132 | 9.56 | $2.42 \times 10^{-16}$ | $R^2$ | 0.188 |
| GDP | $7.18 \times 10^{-7}$ | $4.04 \times 10^{-7}$ | 1.78 | 0.0778 | $N$ | 120 |
| OLD | 0.0027 | 0.00076 | 3.55 | 0.000557 | Cross-correlation | -0.4561 |

**Figure 3:**
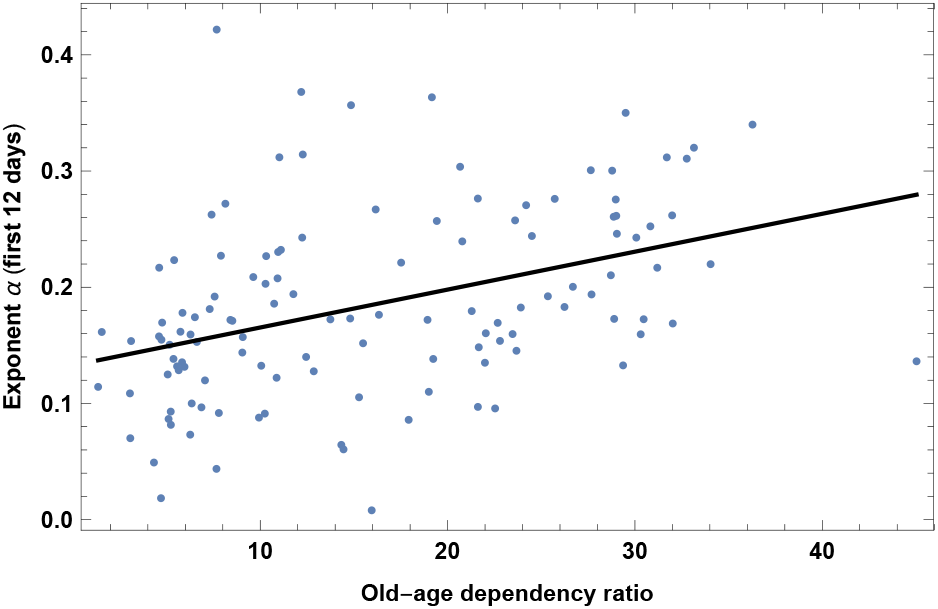
Exponent *α* for each country vs. old-age dependency ratio, as defined in the text. We show the data points and the best-fit for the linear interpolation.

Note that a similar variable is life expectancy (which we analyze later); other variables are also highly correlated, such as median age and child dependency ratio, which we do not show here. In analogy to the previous interpretation, data indicate that a younger population, including countries with high percentage of children, is more immune to COVID-19, or less contagious.

### 3. Life expectancy

This dataset is for year 2016. It has high correlation with old-age dependency ratio. It also has high correlations with other datasets in [13] that we do not show here, such as median age and child dependency ratio (the ratio between under-19-year-olds and 20-to-69-year-olds). Results are shown in fig. 4 and Table IV.

**Table IV:**
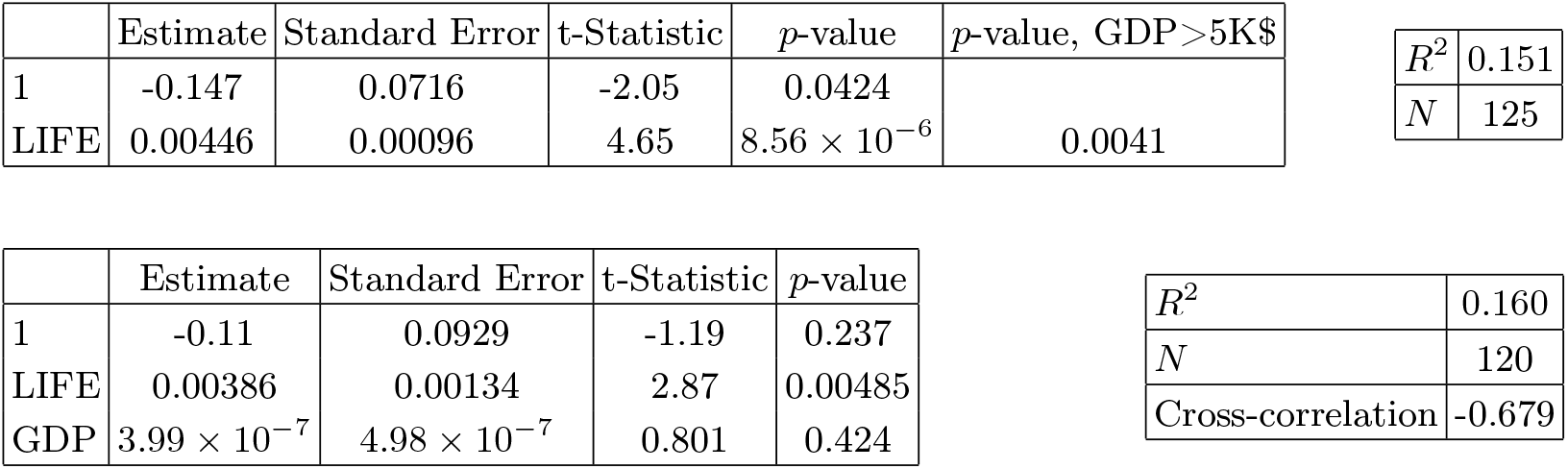
In the left top panel: best-estimate, standard error (*σ*), t-statistic and *p*-value for the parameters of the linear interpolation, for correlation of *α* with life expectancy, LIFE. We also show the *p*-value, excluding countries below 5 thousand $ GDP per capita. In the left bottom panel: same quantities for correlation of *α* with LIFE and GDP per capita. In the right panels: *R*^2^ for the best-estimate and number of countries *N*. We also show the correlation coefficient between the 2 variables in the two-variable fit.

| | Estimate | Standard Error | t-Statistic | $p$-value | $p$-value, GDP>5K\$ |
| --- | --- | --- | --- | --- | --- |
| 1 | -0.147 | 0.0716 | -2.05 | 0.0424 |  |
| LIFE | 0.00446 | 0.00096 | 4.65 | $8.56 \times 10^{-6}$ | 0.0041 |

|  |  |
| --- | --- |
| $R^2$ | 0.151 |
| $N$ | 125 |

| | Estimate | Standard Error | t-Statistic | $p$-value |
| --- | --- | --- | --- | --- |
| 1 | -0.11 | 0.0929 | -1.19 | 0.237 |
| LIFE | 0.00386 | 0.00134 | 2.87 | 0.00485 |
| GDP | $3.99 \times 10^{-7}$ | $4.98 \times 10^{-7}$ | 0.801 | 0.424 |

Table IV: In the left top panel: best-estimate, standard error ( $\sigma$ ), t-statistic and $p$ -value for the parameters of the linear interpolation, for correlation of $\alpha$ with life expectancy, LIFE.
|  |  |
| --- | --- |
| $R^2$ | 0.160 |
| $N$ | 120 |
| Cross-correlation | -0.679 |

**Figure 4:**
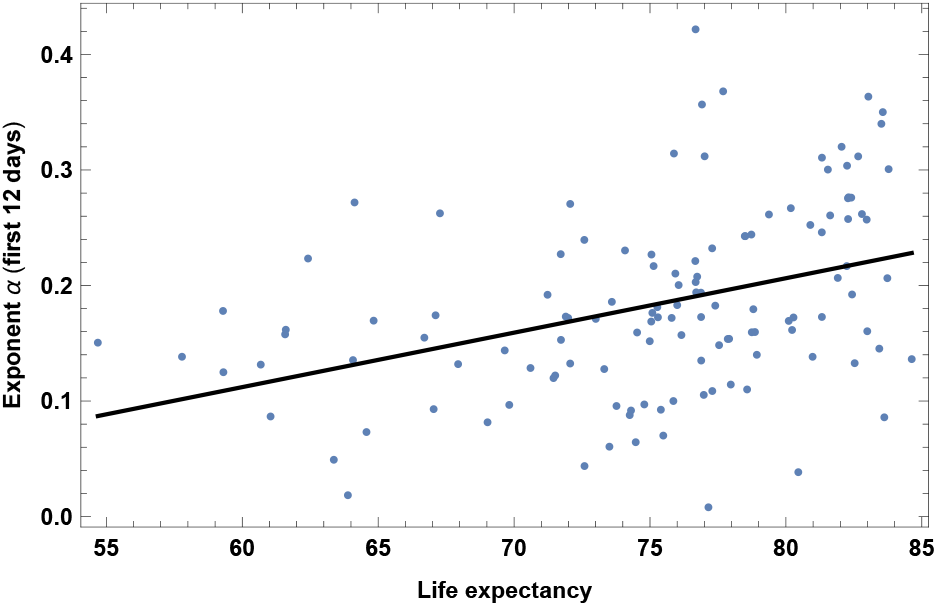
Exponent *α* for each country vs. life expectancy. We show the data points and the best-fit for the linear interpolation.

### 4. International tourism: number of arrivals

The dataset is for year 2016. Results are shown in fig. 5 and Table V. As expected, more tourists correlate with higher speed of contagion. This is in agreement with [2, 10], that found air travel to be an important factor, which will appear here as the number of tourists as well as a correlation with GDP.

**Figure 5:**
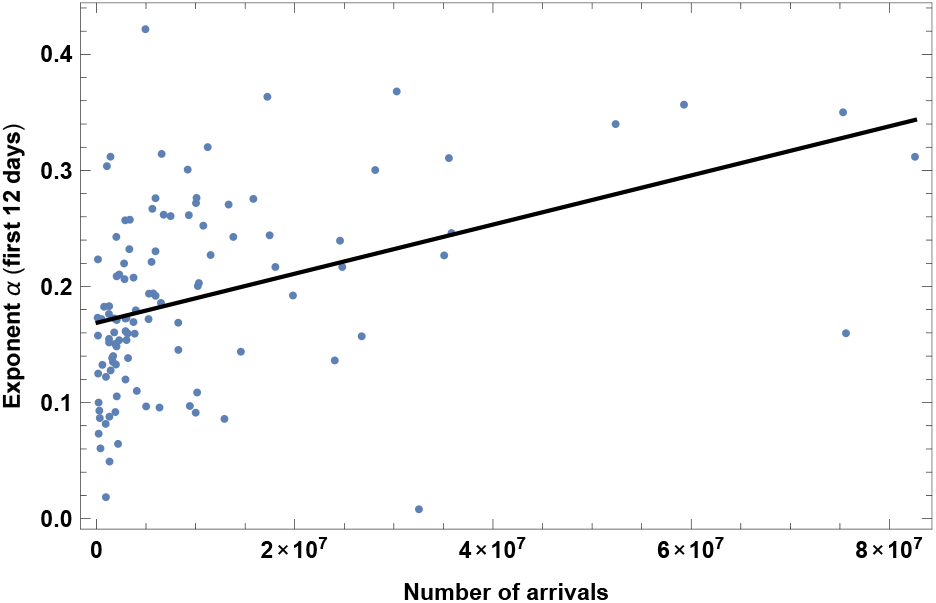
Exponent *α* for each country vs. number of tourist arrivals. We show the data points and the best-fit for the linear interpolation.

### 5. Starting date of the epidemic

This refers to the day *d*_*i*_ chosen as a starting point, counted from December 31st 2019. Results are shown in fig. 6 and Table VI, which shows that earlier contagion is correlated with faster contagion. One possible interpretation is that countries which are affected later are already more aware of the pandemic and therefore have a larger amount of social distancing, which makes the growth rate smaller. Another possible interpretation is that there is some other underlying factor that protects against contagion, and therefore epidemics spreads both later *and* slower.

**Table V:** In the left top panel: best-estimate, standard error (*σ*), t-statistic and *p*-value for the parameters of the linear interpolation, for correlation of *α* with number of tourist arrivals, ARR. We also show the *p*-value, excluding countries below 5 thousand $ GDP per capita. In the left bottom panel: same quantities for correlation of *α* with ARR and GDP per capita. In the right panels: *R*^2^ for the best-estimate and number of countries *N*. We also show the correlation coefficient between the 2 variables in the two-variable fit.

| | Estimate | Standard Error | t-Statistic | $p$-value | $p$-value, GDP>5K\$ | $R^2$ | $N$ |
| --- | --- | --- | --- | --- | --- | --- | --- |
| 1 | 0.168 | 0.00846 | 19.9 | $7.38 \times 10^{-38}$ | | 0.169 | |
| ARR | $2.13 \times 10^{-9}$ | $4.55 \times 10^{-10}$ | 4.69 | $8.12 \times 10^{-6}$ | 0.0002 | | 110 |

Table V: In the left top panel: best-estimate, standard error ( $\sigma$ ), t-statistic and $p$ -value for the parameters of the linear interpolation, for correlation of $\alpha$ with number of tourist arrivals, ARR.
| | Estimate | Standard Error | t-Statistic | $p$-value | $R^2$ | $N$ |
| --- | --- | --- | --- | --- | --- | --- |
| 1 | 0.146 | 0.0116 | 12.6 | $1.48 \times 10^{-22}$ | 0.226 | |
| GDP | $1.11 \times 10^{-6}$ | $4.02 \times 10^{-7}$ | 2.76 | 0.00691 | | 107 |
| ARR | $1.77 \times 10^{-9}$ | $4.68 \times 10^{-10}$ | 3.78 | 0.000265 | Cross-correlation | -0.288 |

**Figure 6:**
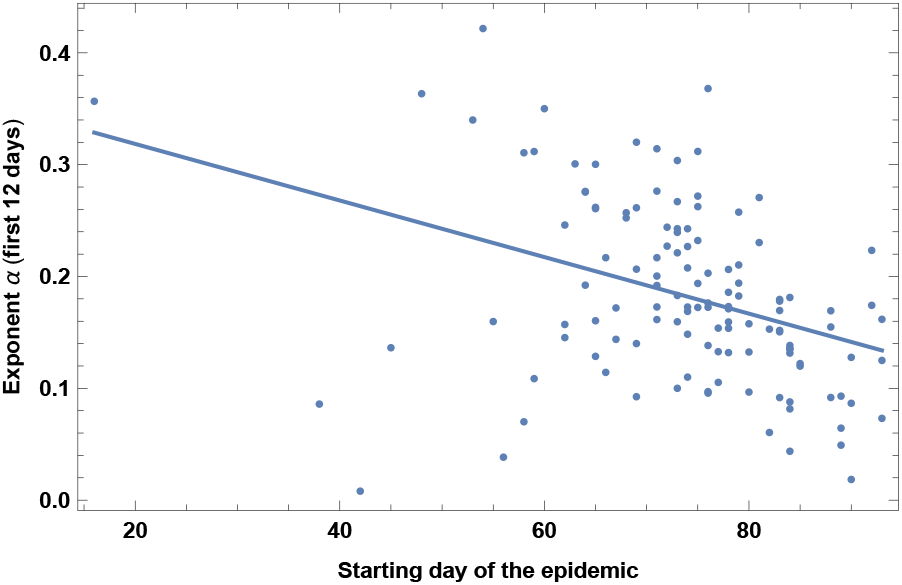
Exponent *α* for each country vs. starting date of the analysis of the epidemic, DATE, defined as the day when the positive cases reached *N* = 30. Days are counted from Dec 31st 2019. We show the data points and the best-fit for the linear interpolation.

### 6. Greeting habits

A relevant variable is the level of contact in greeting habits in each country. We have subdivided the countries in groups according to the physical contact in greeting habits; information has been taken from [28].

1. No or little physical contact, bowing. In this group we have: Bangladesh, Cambodia, Japan, Korea South, Sri Lanka, Thailand.
2. Handshaking between man-man and woman-woman. No or little contact man-woman. In this group we have: India, Indonesia, Niger, Senegal, Singapore, Togo, Vietnam, Zambia
3. Handshaking. In this group we have: Australia, Austria, Bulgaria, Burkina Faso, Canada, China, Estonia, Finland, Germany, Ghana, Madagascar, Malaysia, Mali, Malta, New Zealand, Norway, Philippines, Rwanda, Sweden, Taiwan, Uganda, Kingdom United, States United.
4. Handshaking, plus kissing among friends and relatives, but only man-man and woman-woman. No or little contact man-woman. In this group we have: Afghanistan, Azerbaijan, Bahrain, Belarus, Brunei, Egypt, Guinea, Jordan, Kuwait, Kyrgyzstan, Oman, Pakistan, Qatar, Arabia Saudi, Arab Emirates United, Uzbekistan.
5. Handshaking, plus kissing among friends and relatives. In this group we have: Albania, Algeria, Argentina, Armenia, Belgium, Bolivia, and Bosnia Herzegovina, Cameroon, Chile, Colombia, Costa Rica, Côte d’Ivoire, Croatia, Cuba, Cyprus, Czech Republic, Denmark, Dominican Republic, Ecuador, El Salvador, France, Georgia, Greece, Guatemala, Honduras, Hungary, Iran, Iraq, Ireland, Israel, Italy, Jamaica, Kazakhstan, Kenya, Kosovo, Latvia, Lebanon, Lithuania, Luxembourg, Macedonia, Mauritius, Mexico, Moldova, Montenegro, Morocco, Netherlands, Panama, Paraguay, Peru, Poland, Portugal, Puerto Rico, Romania, Russia, Serbia, Slovakia, Slovenia, Africa South, Switzerland, Trinidad and Tobago, Tunisia, Turkey, Ukraine, Uruguay, Venezuela.
6. Handshaking and kissing. In this group we have: Andorra, Brazil, Spain.

We have arbitrarily assigned a variable, named GRE, from 0 to 1 to each group, namely GRE = 0, 0.25, 0.5, 0.4, 0.8 and 1, respectively. We have chosen a ratio of 2 between group 2 and 3 and between group 4 and 5, based on the fact that the only difference is that about half of the possible interactions (men-women) are without contact. Results are shown in Fig. 7 and Table VII.

**Figure 7:**
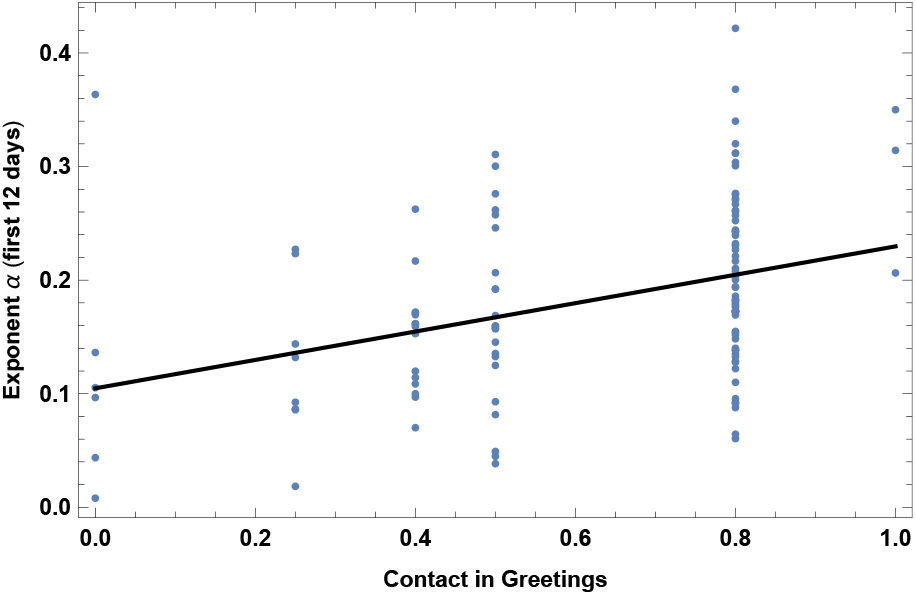
Exponent *α* for each country vs. level of contact in greeting habits, *GRE*, as defined in the text. We show the data points and the best-fit for the linear interpolation.

Note also that an outlier is visible in the plot, which corresponds to South Korea. The early outbreak of the disease in this particular case was strongly affected by the Shincheonji Church, which included mass prayer and worship sessions. By excluding South Korea from the dataset one finds an even larger significance, *p*-value= 6.4 · 10^−8^ and *R*^2^ *≈* 0.23.

### 7. Lung cancer death rates

This dataset refers to year 2002. Results are shown in Fig. 8 and Table VIII. Such results are interesting and could be interpreted a priori in two ways. A first interpretation is that COVID-19 contagion might correlate to lung cancer, simply due to the fact that lung cancer is more prevalent in countries with more old people. Such a simplistic interpretation is somehow contradicted by the case of generic cancer death rates, discussed in section III, which is indeed less significant than lung cancer. A better interpretation is therefore that lung cancer may be a specific risk factor for COVID-19 contagion. This is supported also by the observation of high rates of lung cancer in COVID-19 patients [14].

**Table VI:**
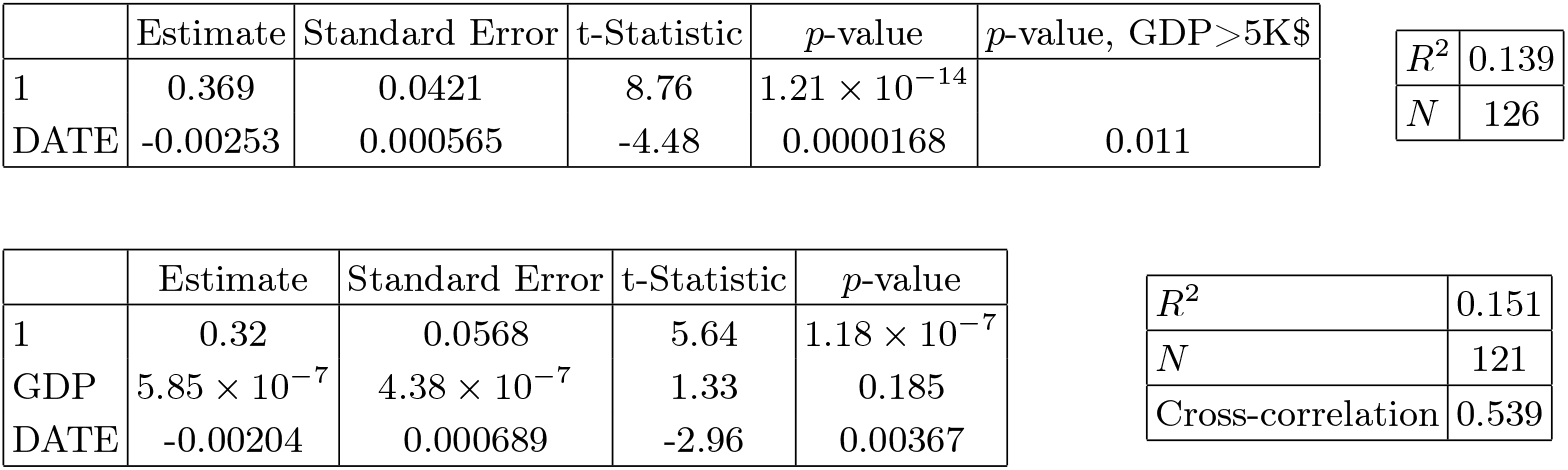
In the left top panel: best-estimate, standard error (*σ*), t-statistic and *p*-value for the parameters of the linear interpolation, for correlation of *α* with vs. starting date of the analysis of the epidemic, DATE, as defined in the text. We also show the *p*-value, excluding countries below 5 thousand $ GDP per capita. In the left bottom panel: same quantities for correlation of *α* with DATE and GDP per capita. In the right panels: *R*^2^ for the best-estimate and number of countries *N*. We also show the correlation coefficient between the 2 variables in the two-variable fit.

**Table VII:**
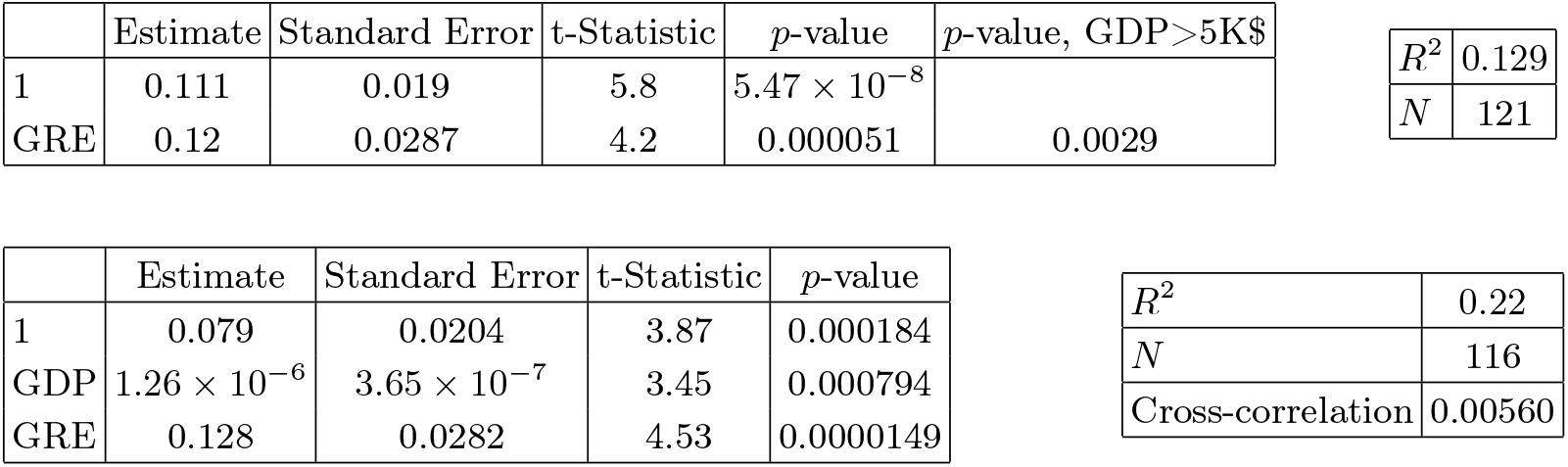
In the left top panel: best-estimate, standard error (*σ*), t-statistic and *p*-value for the parameters of the linear interpolation, for correlation of *α* with level of contact in greeting habits, GRE, as defined in the text. We also show the *p*-value, excluding countries below 5 thousand $ GDP per capita. In the left bottom panel: same quantities for correlation of *α* with GRE and GDP per capita. In the right panels: *R*^2^ for the best-estimate and number of countries *N*. We also show the correlation coefficient between the 2 variables in the two-variable fit.

| | Estimate | Standard Error | t-Statistic | $p\text{-value}$ | $p\text{-value, GDP} > 5K\$$ |
| --- | --- | --- | --- | --- | --- |
| 1 | 0.111 | 0.019 | 5.8 | $5.47 \times 10^{-8}$ | |
| $GRE$ | 0.12 | 0.0287 | 4.2 | 0.000051 | 0.0029 |

|  |  |
| --- | --- |
| $R^2$ | 0.129 |
| $N$ | 121 |

| | Estimate | Standard Error | t-Statistic | $p\text{-value}$ |
| --- | --- | --- | --- | --- |
| 1 | 0.079 | 0.0204 | 3.87 | 0.000184 |
| $GDP$ | $1.26 \times 10^{-6}$ | $3.65 \times 10^{-7}$ | 3.45 | 0.000794 |
| $GRE$ | 0.128 | 0.0282 | 4.53 | 0.0000149 |

Table VII: In the left top panel: best-estimate, standard error ( $\sigma$ ), t-statistic and $p\text{-value}$ for the parameters of the linear interpolation, for correlation of $\alpha$ with level of contact in greeting habits, $GRE$ , as defined in the text.
|  |  |
| --- | --- |
| $R^2$ | 0.22 |
| $N$ | 116 |
| Cross-correlation | 0.00560 |

**Figure 8:**
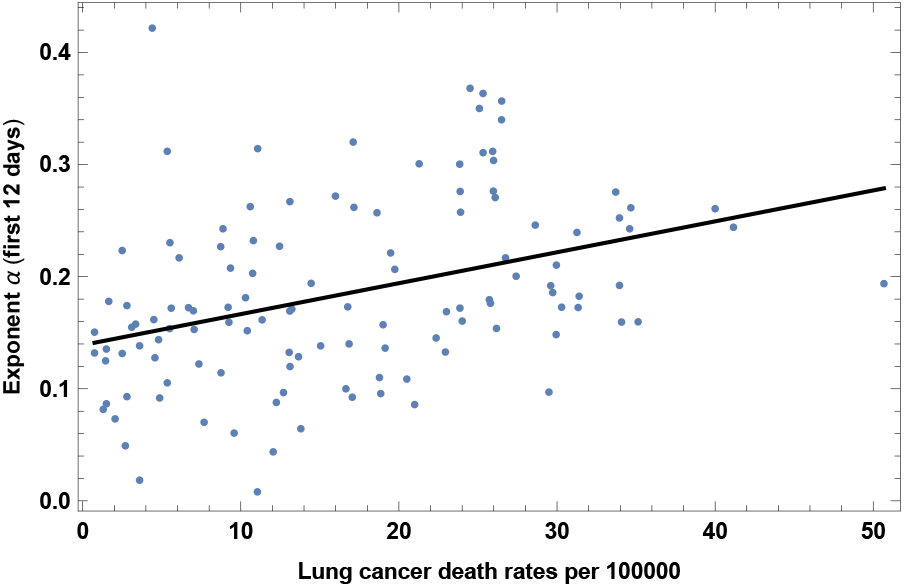
Exponent *α* for each country vs. lung cancer death rates. We show the data points and the best-fit for the linear interpolation.

### 8. Obesity in males

This refers to the prevalence of obesity in adult *males*, measured in 2014. Results are shown in fig. 9 and Table IX. Note that this effect is mostly due to the difference between very poor countries and the rest of the world; indeed this becomes non-significant when excluding countries below 5K$ GDP per capita. Note also that obesity in *females* instead is *not* correlated with growth rate of COVID-19 contagion in our sample. See also [15] for increased risk of severe COVID-19 symptoms for obese patients.

**Figure 9:**
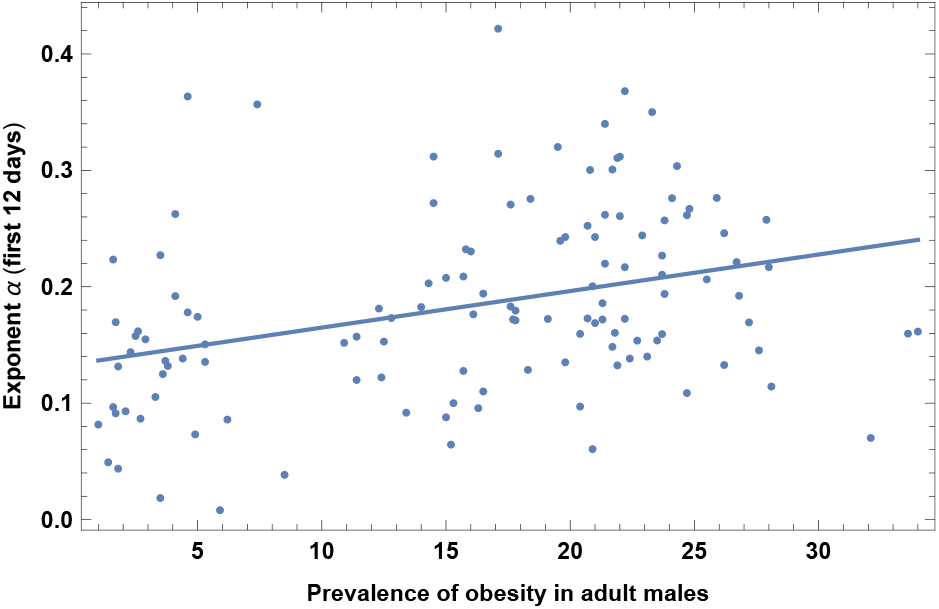
Exponent *α* for each country vs. prevalence of obesity in adult males. We show the data points and the best-fit for the linear interpolation.

### 9. Urbanization

This is the share of population living in urban areas, collected in year 2017. Results are shown in Fig. 10 and Table X. This is an expected correlation, in agreement with [19, 20].

**Table VIII:** In the left top panel: best-estimate, standard error (*σ*), t-statistic and *p*-value for the parameters of the linear interpolation, for correlation of *α* with lung cancer death rates, LUNG. We also show the *p*-value, excluding countries below 5 thousand $ GDP per capita. In the left bottom panel: same quantities for correlation of *α* with LUNG and GDP per capita. In the right panels: *R*^2^ for the best-estimate and number of countries *N*. We also show the correlation coefficient between the 2 variables in the two-variable fit.

| | Estimate | Standard Error | t-Statistic | $p$-value | $p$-value, GDP>5K\$ | $R^2$ | $N$ |
| --- | --- | --- | --- | --- | --- | --- | --- |
| 1 | 0.143 | 0.0121 | 11.8 | $9.96 \times 10^{-22}$ | | 0.127 | 121 |
| LUNG | 0.00159 | 0.000381 | 4.17 | 0.0000572 | 0.024 |  |  |

Table VIII: In the left top panel: best-estimate, standard error ( $\sigma$ ), t-statistic and $p$ -value for the parameters of the linear interpolation, for correlation of $\alpha$ with lung cancer death rates, LUNG.
| | Estimate | Standard Error | t-Statistic | $p$-value | $R^2$ | $N$ |
| --- | --- | --- | --- | --- | --- | --- |
| 1 | 0.133 | 0.013 | 10.2 | $7.48 \times 10^{-18}$ | 0.164 | 118 |
| GDP | $8.93 \times 10^{-7}$ | $4.02 \times 10^{-7}$ | 2.22 | 0.0282 | Cross-correlation | -0.403 |
| LUNG | 0.00122 | 0.000416 | 2.94 | 0.00396 |  |  |

**Table IX:**
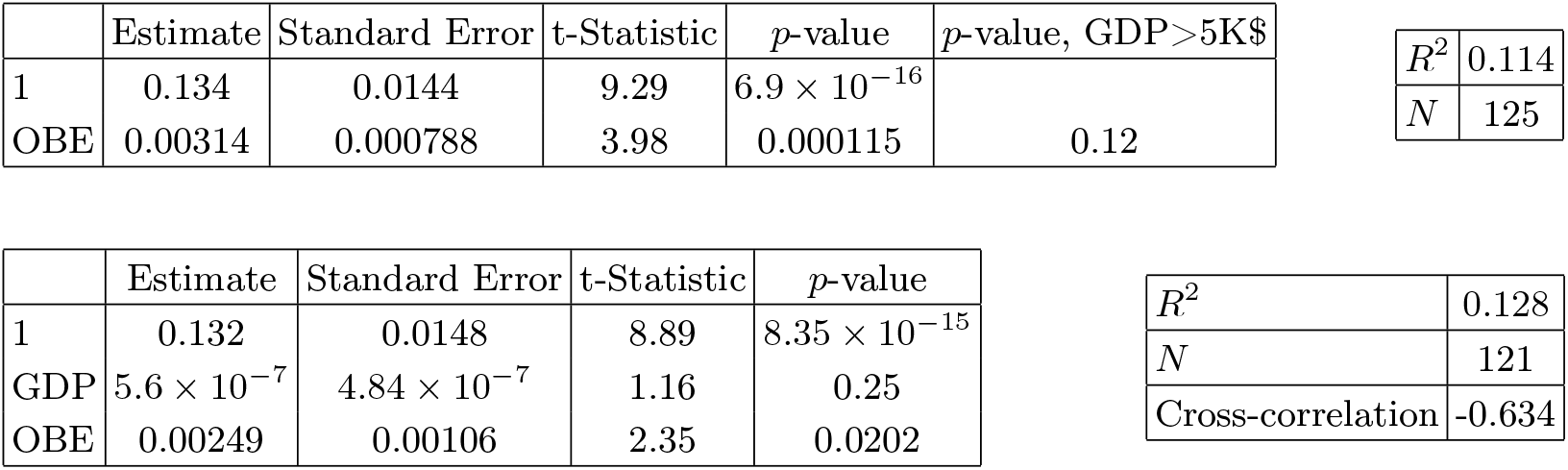
In the left top panel: best-estimate, standard error (*σ*), t-statistic and *p*-value for the parameters of the linear interpolation, for correlation of *α* with prevalence of obesity in adult males (OBE). We also show the *p*-value, excluding countries below 5 thousand $ GDP per capita. In the left bottom panel: same quantities for correlation of *α* with OBE and GDP per capita. In the right panels: *R*^2^ for the best-estimate and number of countries *N*. We also show the correlation coefficient between the 2 variables in the two-variable fit.

| | Estimate | Standard Error | t-Statistic | $p$-value | $p$-value, GDP>5K\$ |
| --- | --- | --- | --- | --- | --- |
| 1 | 0.134 | 0.0144 | 9.29 | $6.9 \times 10^{-16}$ | |
| OBE | 0.00314 | 0.000788 | 3.98 | 0.000115 | 0.12 |

|  |  |
| --- | --- |
| $R^2$ | 0.114 |
| $N$ | 125 |

| | Estimate | Standard Error | t-Statistic | $p$-value |
| --- | --- | --- | --- | --- |
| 1 | 0.132 | 0.0148 | 8.89 | $8.35 \times 10^{-15}$ |
| GDP | $5.6 \times 10^{-7}$ | $4.84 \times 10^{-7}$ | 1.16 | 0.25 |
| OBE | 0.00249 | 0.00106 | 2.35 | 0.0202 |

Table IX: In the left top panel: best-estimate, standard error ( $\sigma$ ), t-statistic and $p$ -value for the parameters of the linear interpolation, for correlation of $\alpha$ with prevalence of obesity in adult males (OBE).
|  |  |
| --- | --- |
| $R^2$ | 0.128 |
| $N$ | 121 |
| Cross-correlation | -0.634 |

**Figure 10:**
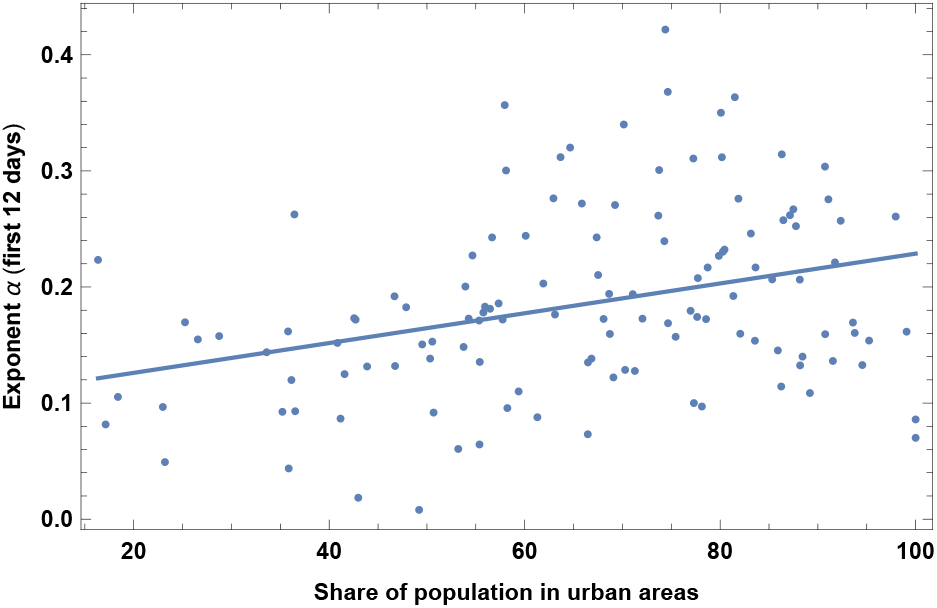
Exponent *α* for each country vs. share of population in urban areas. We show the data points and the best-fit for the linear interpolation.

### 10. Alcohol consumption

This dataset refers to year 2016. Results are shown in Fig. 11 and Table XI. Note that this variable is highly correlated with old-age dependency ratio, as discussed in section V. While the correlation with alcohol consumption may be simply due to correlation with other variables, such as old-age dependency ratio, this finding deserves anyway more research, to assess whether it may be at least partially due to the deleterious effects of alcohol on the immune system.

**Table X:**
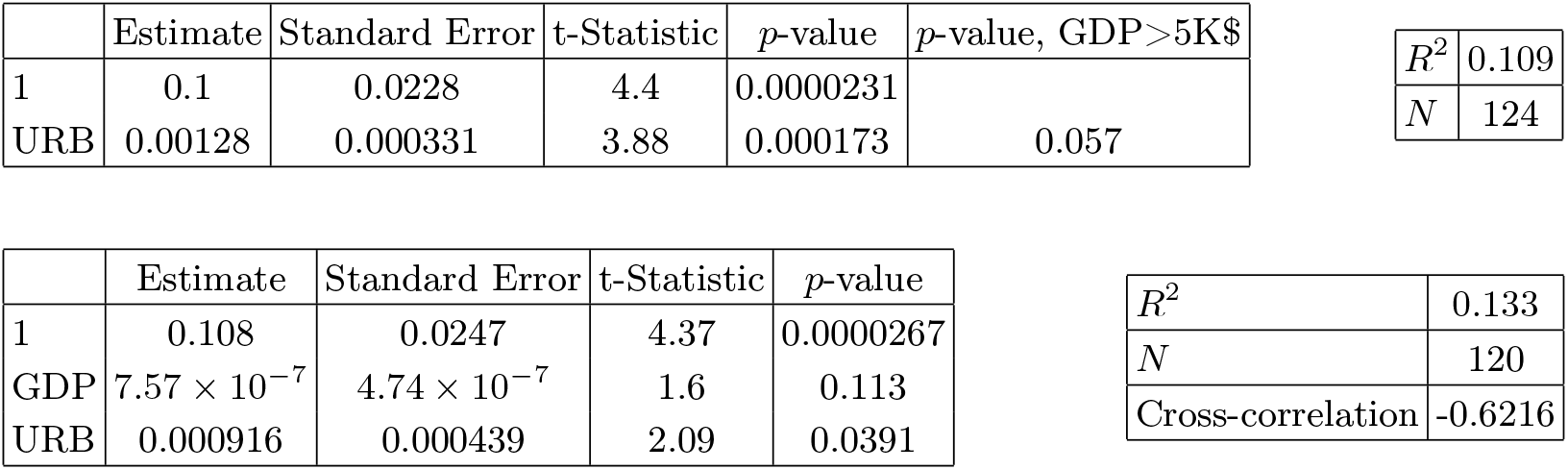
In the left top panel: best-estimate, standard error (*σ*), t-statistic and *p*-value for the parameters of the linear interpolation, for correlation of *α* with share of population in urban areas, URB, as defined in the text. We also show the *p*-value, excluding countries below 5 thousand $ GDP per capita. In the left bottom panel: same quantities for correlation of *α* with URB and GDP per capita. In the right panels: *R*^2^ for the best-estimate and number of countries *N*. We also show the correlation coefficient between the 2 variables in the two-variable fit.

| | Estimate | Standard Error | t-Statistic | $p$-value | $p$-value, GDP>5K\$ |
| --- | --- | --- | --- | --- | --- |
| 1 | 0.1 | 0.0228 | 4.4 | 0.0000231 |  |
| URB | 0.00128 | 0.000331 | 3.88 | 0.000173 | 0.057 |

|  |  |
| --- | --- |
| $R^2$ | 0.109 |
| $N$ | 124 |

| | Estimate | Standard Error | t-Statistic | $p$-value |
| --- | --- | --- | --- | --- |
| 1 | 0.108 | 0.0247 | 4.37 | 0.0000267 |
| GDP | $7.57 \times 10^{-7}$ | $4.74 \times 10^{-7}$ | 1.6 | 0.113 |
| URB | 0.000916 | 0.000439 | 2.09 | 0.0391 |

Table X: In the left top panel: best-estimate, standard error ( $\sigma$ ), t-statistic and $p$ -value for the parameters of the linear interpolation, for correlation of $\alpha$ with share of population in urban areas, URB, as defined in the text.
|  |  |
| --- | --- |
| $R^2$ | 0.133 |
| $N$ | 120 |
| Cross-correlation | -0.6216 |

**Table XI:** In the left top panel: best-estimate, standard error (*σ*), t-statistic and *p*-value for the parameters of the linear interpolation, for correlation of *α* with alcohol consumption (ALCO). We also show the *p*-value, excluding countries below 5 thousand $ GDP per capita. In the left bottom panel: same quantities for correlation of *α* with ALCO and GDP per capita. In the right panels: *R*^2^ for the best-estimate and number of countries *N*. We also show the correlation coefficient between the 2 variables in the two-variable fit.

| | Estimate | Standard Error | t-Statistic | $p$-value | $p$-value, GDP>5K\$ | | |
| --- | --- | --- | --- | --- | --- | --- | --- |
| 1 | 0.15 | 0.0131 | 11.4 | $7.8 \times 10^{-21}$ | | $R^2$ | 0.076 |
| ALCO | 0.00518 | 0.00164 | 3.17 | 0.00195 | 0.012 | $N$ | 126 |

Table XI: In the left top panel: best-estimate, standard error ( $\sigma$ ), t-statistic and $p$ -value for the parameters of the linear interpolation, for correlation of $\alpha$ with alcohol consumption (ALCO).
| | Estimate | Standard Error | t-Statistic | $p$-value | | |
| --- | --- | --- | --- | --- | --- | --- |
| 1 | 0.134 | 0.0141 | 9.48 | $3.75 \times 10^{-16}$ | $R^2$ | 0.138 |
| GDP | $1.12 \times 10^{-6}$ | $3.86 \times 10^{-7}$ | 2.91 | 0.00437 | $N$ | 120 |
| ALCO | 0.00382 | 0.0017 | 2.25 | 0.0264 | Cross-correlation | -0.286 |

**Figure 11:**
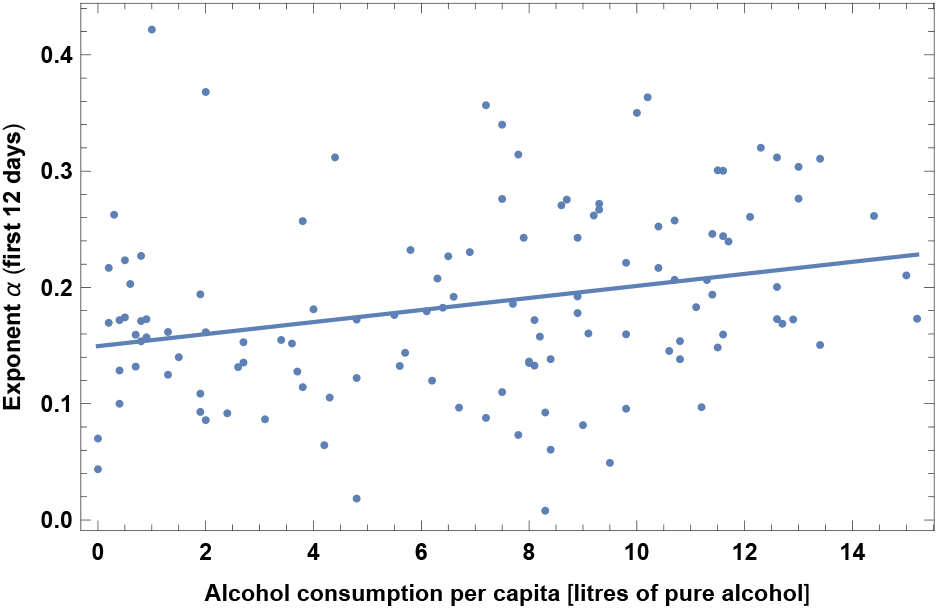
Exponent *α* for each country vs. alcohol consumption. We show the data points and the best-fit for the linear interpolation.

### 11. Smoking

This dataset refers to year 2012. Results are shown in Fig. 12 and Table XII. As expected this variable is highly correlated with lung cancer, as discussed in section V. We find that COVID-19 spreads more rapidly in countries with higher daily smoking prevalence. Note however that this becomes non-significant when excluding countries below 5K$ GDP per capita.

**Table XII:**
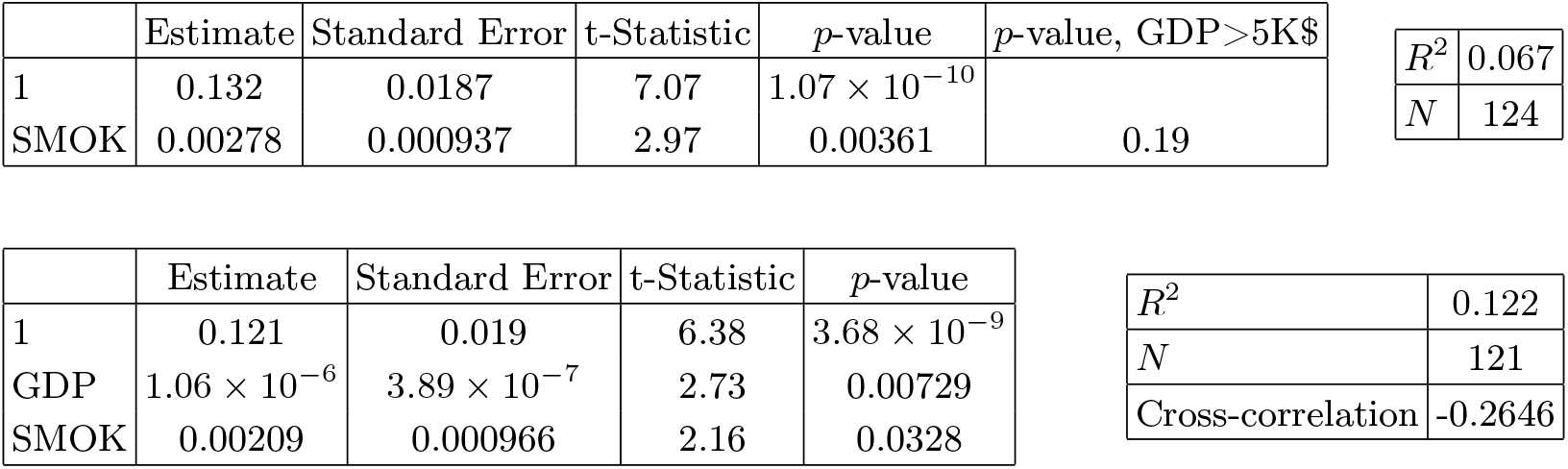
In the left top panel: best-estimate, standard error (*σ*), t-statistic and *p*-value for the parameters of the linear interpolation, for correlation of *α* with daily smoking prevalence (SMOK). We also show the *p*-value, excluding countries below 5 thousand $ GDP per capita. In the left bottom panel: same quantities for correlation of *α* with SMOK and GDP per capita. In the right panels: *R*^2^ for the best-estimate and number of countries *N*. We also show the correlation coefficient between the 2 variables in the two-variable fit.

**Figure 12:**
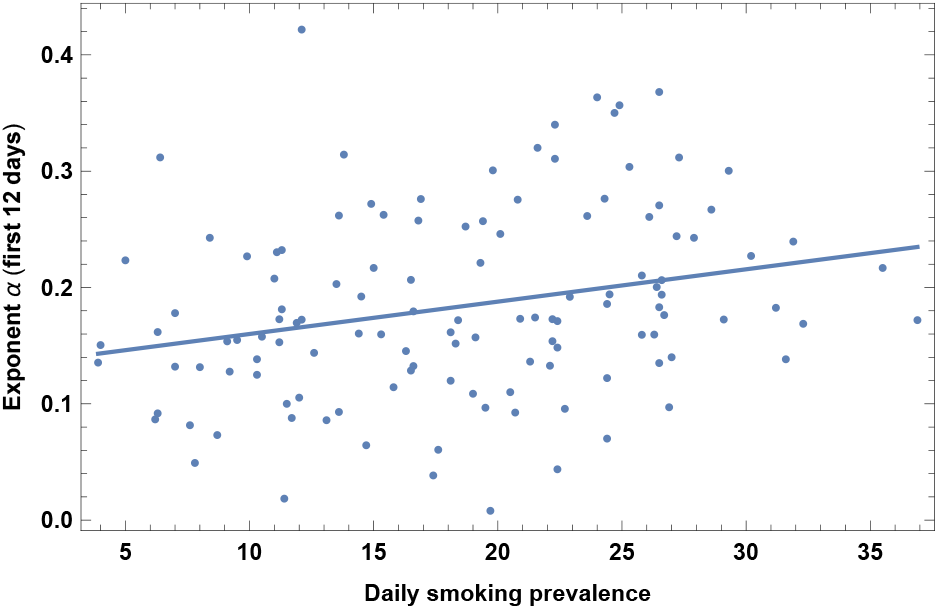
Exponent *α* for each country vs. daily smoking prevalence. We show the data points and the best-fit for the linear interpolation.

Correlation of *α* with smoking thus could be simply due to correlation with other variables or to a bias due to lack of testing in very poor countries. Alternative interpretations are that smoking has negative effects on conditions of lungs that facilitates contagion or that it contributes to increased transmission of virus from hand to mouth [16]. Interestingly, note that our finding is in contrast with claims of a possible protective effect of nicotine and smoking against COVID-19 [17, 18].

### 12. UV index

This is the UV index for the relevant period of time of the epidemic. In particular the UV index has been collected from [21], as a monthly average, and then with a linear interpolation we have used the average value during the 12 days of the epidemic growth, for each country. Results are shown in Fig. 13 and Table XIII. Not surprisingly in section V we will see that such quantity is very highly correlated with T (correlation coefficient 0.93). Note also that here the sample size is smaller (73) than in the case of other variables, so it is not strange that the significance of a correlation with *α* here is not as high as in the case of *α* with Temperature. More research is required to answer more specific questions, for instance whether the virus survives less in an environment with high UV index, or whether a high UV index stimulates vitamin D production that may help the immune system, or both. Very recent results [56] consistently find that indeed SARS-CoV-2 is deactivated efficiently under simulated sunlight in few minutes, due to UVB.

**Table XIII:**
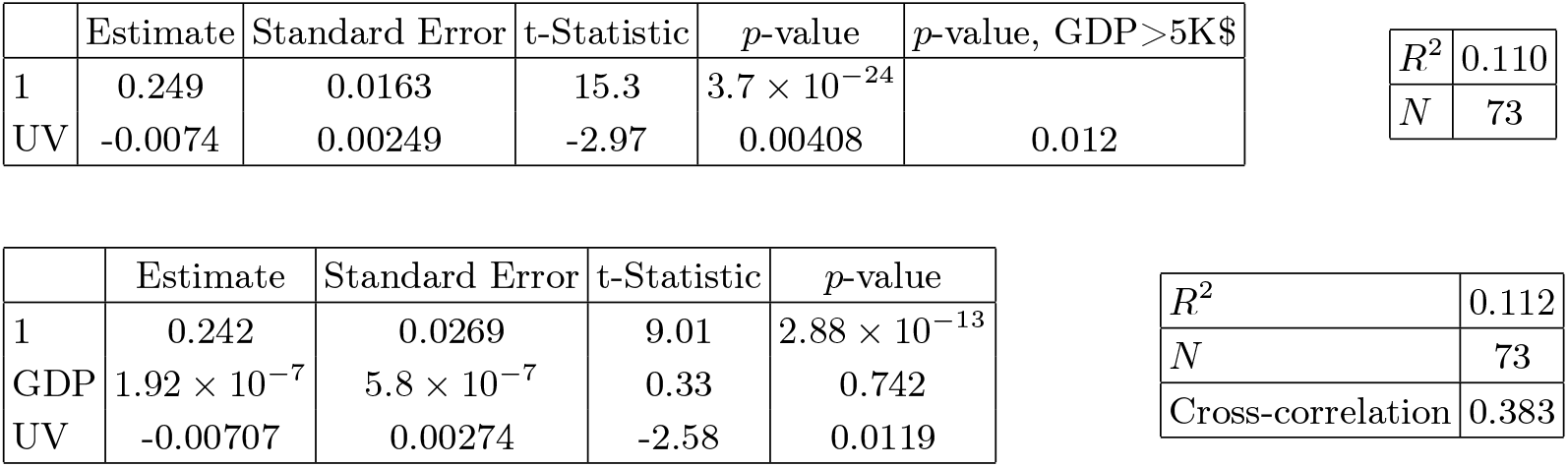
In the left top panel: best-estimate, standard error (*σ*), t-statistic and *p*-value for the parameters of the linear interpolation, for correlation of *α* with UV index for the relevant period of time of the epidemic (UV). We also show the *p*-value, excluding countries below 5 thousand $ GDP per capita. In the left bottom panel: same quantities for correlation of *α* with UV and GDP per capita. In the right panels: *R*^2^ for the best-estimate and number of countries *N*. We also show the correlation coefficient between the 2 variables in the two-variable fit.

| | Estimate | Standard Error | t-Statistic | $p$-value | $p$-value, GDP > 5K\$ |
| --- | --- | --- | --- | --- | --- |
| 1 | 0.249 | 0.0163 | 15.3 | $3.7 \times 10^{-24}$ | |
| UV | -0.0074 | 0.00249 | -2.97 | 0.00408 | 0.012 |

|  |  |
| --- | --- |
| $R^2$ | 0.110 |
| $N$ | 73 |

| | Estimate | Standard Error | t-Statistic | $p$-value |
| --- | --- | --- | --- | --- |
| 1 | 0.242 | 0.0269 | 9.01 | $2.88 \times 10^{-13}$ |
| GDP | $1.92 \times 10^{-7}$ | $5.8 \times 10^{-7}$ | 0.33 | 0.742 |
| UV | -0.00707 | 0.00274 | -2.58 | 0.0119 |

Table XIII: In the left top panel: best-estimate, standard error ( $\sigma$ ), t-statistic and $p$ -value for the parameters of the linear interpolation, for correlation of $\alpha$ with UV index for the relevant period of time of the epidemic (UV).
|  |  |
| --- | --- |
| $R^2$ | 0.112 |
| $N$ | 73 |
| Cross-correlation | 0.383 |

**Figure 13:**
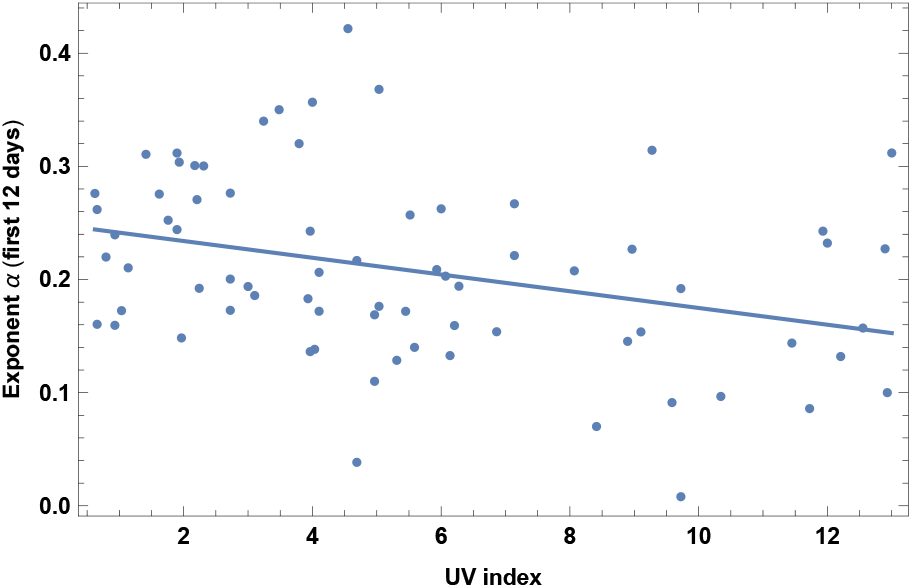
Exponent *α* for each country vs. UV index for the relevant month of the epidemic. We show the data points and the best-fit for the linear interpolation.

### 13. Vitamin D serum concentration

Another relevant variable is the amount of serum Vitamin D. We collected data in the literature for the average annual level of serum Vitamin D and for the seasonal level (D_*s*_). The seasonal level is defined as: the amount during the month of March or during winter for northern hemisphere, *or* during summer for southern hemisphere *or* the annual level for countries with little seasonal variation. The dataset for the annual D was built with the available literature, which is unfortunately quite inhomogeneous as discussed in Appendix A. For many countries several studies with quite different values were found and in this case we have collected the mean and the standard error and a weighted average has been performed. The countries included in this dataset are 50, as specified in Appendix. The dataset for the seasonal levels is more restricted, since the relative literature is less complete, and we have included 42 countries.

Results are shown in Fig. 14 and Table XXXIV for the annual levels and in Fig. 14 and Table XXXIV for the seasonal levels. Note however that such results are preliminary and based on inhomogeneous data, and have to be confirmed on a larger sample.

**Figure 14:**
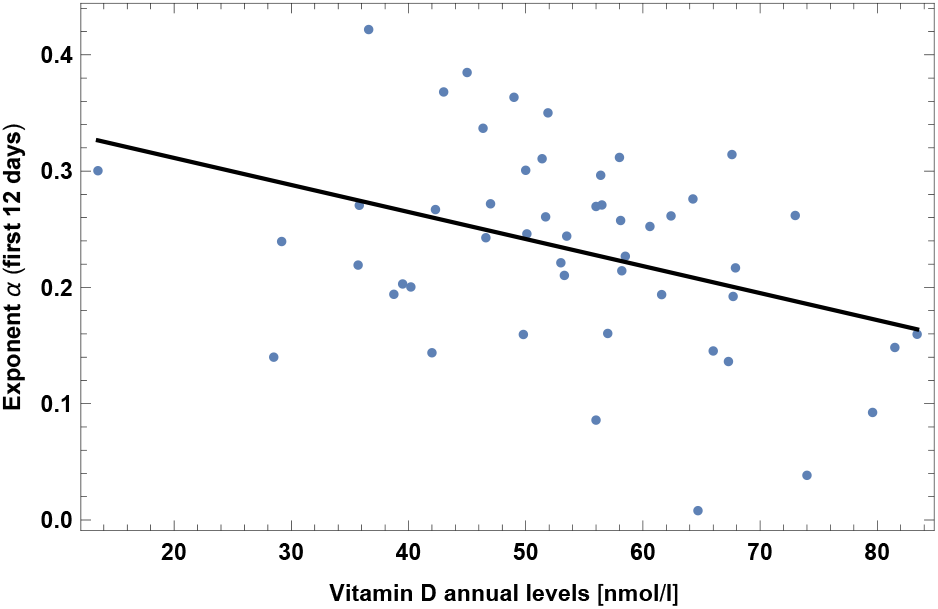
Exponent *α* for each country vs. annual levels of vitamin *D*, for the relevant period of time, as defined in the text, for the base set of 42 countries. We show the data points and the best-fit for the linear interpolation.

**Figure 15:**
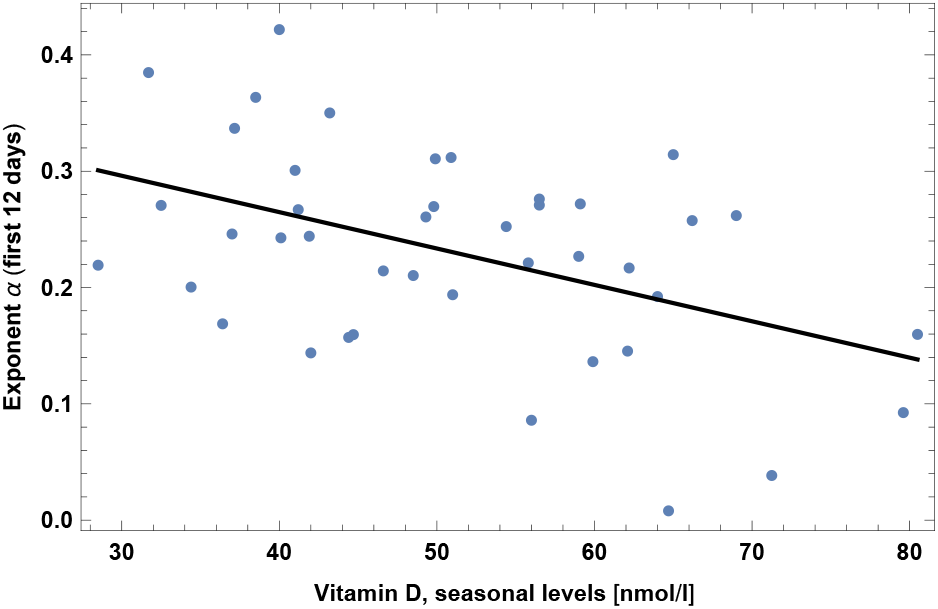
Exponent *α* for each country vs. seasonal levels of vitamin D, for the relevant period of time, as defined in the text, for the base set of 42 countries. We show the data points and the best-fit for the linear interpolation.

Interestingly, in section V we will see that D is *not* highly correlated with T or UV index, as one naively could expect, due to different food consumption in different countries. A slightly higher correlation, as it should be, is present between T and D_*s*_. Note that our results are in agreement with the fact that increased vitamin D levels have been proposed to have a protective effect against COVID-19 [45–47].

Table XIV: In the left top panel: best-estimate, standard error (*σ*), t-statistic and *p*-value for the parameters of the linear interpolation, for correlation of *α* with mean annual levels of vitamin D (variable name: D). We also show the *p*-value, excluding countries below 5 thousand $ GDP per capita. In the left bottom panel: same quantities for correlation of *α* with D and GDP per capita. In the right panels: *R*^2^ for the best-estimate and number of countries *N*. We also show the correlation coefficient between the 2 variables in the two-variable fit.

### 14. CO_2_ Emissions

This is the data for year 2017. We have also checked that this has very high correlation with SO emissions (about 0.9 correlation coefficient). We show here only the case for CO_2_, but the reader should keep in mind that a very similar result applies also to SO emissions. Note also that this is expected to have a high correlation with the number of international tourist arrivals, as we will show in section V. Results are shown in Fig. 16 and Table XVI.

**Table XIV:**
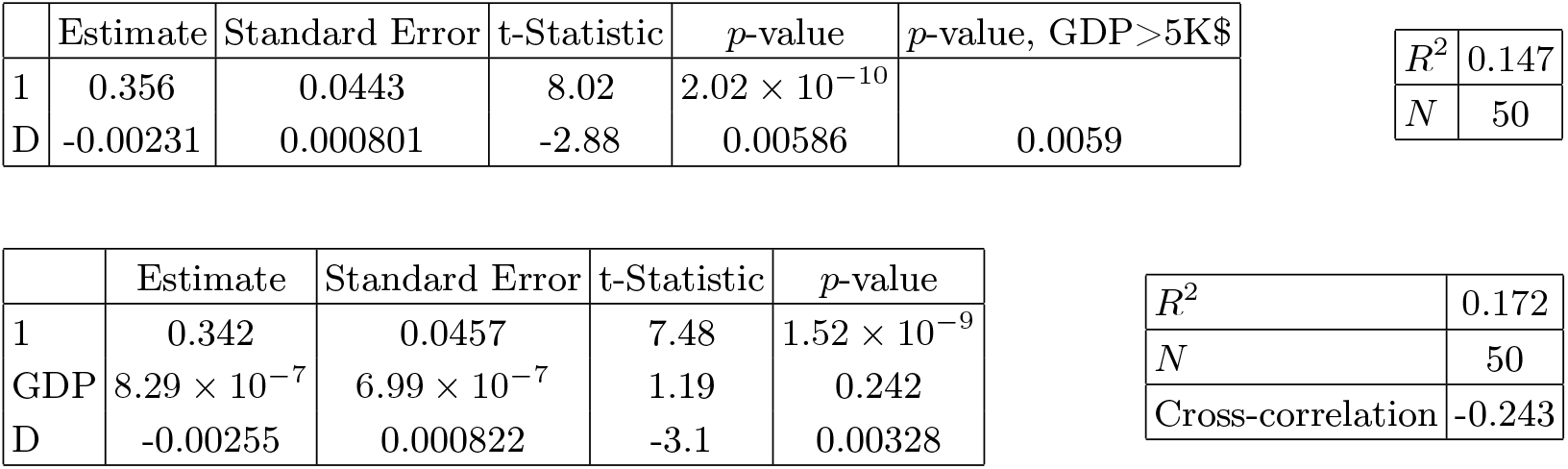
In the left top panel: best-estimate, standard error (*σ*), t-statistic and *p*-value for the parameters of the linear interpolation, for correlation of *α* with seasonal levels of vitamin D (variable name: D_*s*_). We also show the *p*-value, excluding countries below 5 thousand $ GDP per capita. In the left bottom panel: same quantities for correlation of *α* with D_*s*_ and GDP per capita. In the right panels: *R*^2^ for the best-estimate and number of countries *N*. We also show the correlation coefficient between the 2 variables in the two-variable fit.

| | Estimate | Standard Error | t-Statistic | $p$-value | $p$-value, GDP>5K\$ |
| --- | --- | --- | --- | --- | --- |
| 1 | 0.356 | 0.0443 | 8.02 | $2.02 \times 10^{-10}$ | |
| D | -0.00231 | 0.000801 | -2.88 | 0.00586 | 0.0059 |

|  |  |
| --- | --- |
| $R^2$ | 0.147 |
| $N$ | 50 |

| | Estimate | Standard Error | t-Statistic | $p$-value |
| --- | --- | --- | --- | --- |
| 1 | 0.342 | 0.0457 | 7.48 | $1.52 \times 10^{-9}$ |
| GDP | $8.29 \times 10^{-7}$ | $6.99 \times 10^{-7}$ | 1.19 | 0.242 |
| D | -0.00255 | 0.000822 | -3.1 | 0.00328 |

Table XIV: In the left top panel: best-estimate, standard error ( $\sigma$ ), t-statistic and $p$ -value for the parameters of the linear interpolation, for correlation of $\alpha$ with mean annual levels of vitamin D (variable name: D).
|  |  |
| --- | --- |
| $R^2$ | 0.172 |
| $N$ | 50 |
| Cross-correlation | -0.243 |

**Table XV:**
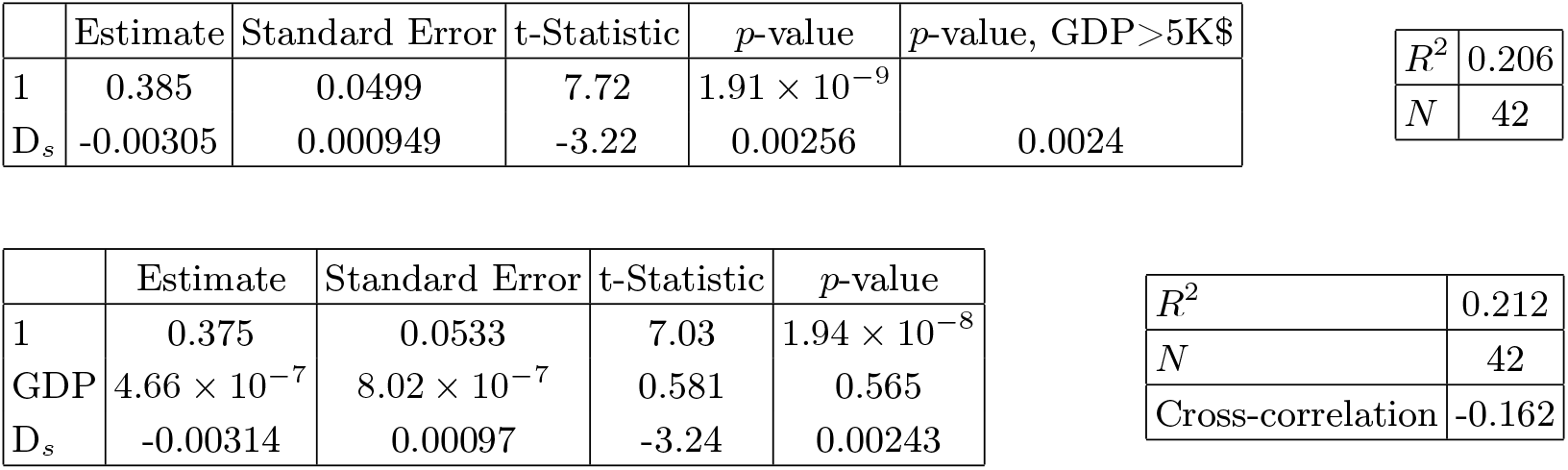
In the left top panel: best-estimate, standard error (*σ*), t-statistic and *p*-value for the parameters of the linear interpolation, for correlation of *α* with seasonal levels of vitamin D (variable name: D_*s*_). We also show the *p*-value, excluding countries below 5 thousand $ GDP per capita. In the left bottom panel: same quantities for correlation of *α* with D_*s*_ and GDP per capita. In the right panels: *R*^2^ for the best-estimate and number of countries *N*. We also show the correlation coefficient between the 2 variables in the two-variable fit.

| | Estimate | Standard Error | t-Statistic | $p$-value | $p$-value, GDP>5K\$ |
| --- | --- | --- | --- | --- | --- |
| 1 | 0.385 | 0.0499 | 7.72 | $1.91 \times 10^{-9}$ | |
| $D_s$ | -0.00305 | 0.000949 | -3.22 | 0.00256 | 0.0024 |

|  |  |
| --- | --- |
| $R^2$ | 0.206 |
| $N$ | 42 |

| | Estimate | Standard Error | t-Statistic | $p$-value |
| --- | --- | --- | --- | --- |
| 1 | 0.375 | 0.0533 | 7.03 | $1.94 \times 10^{-8}$ |
| GDP | $4.66 \times 10^{-7}$ | $8.02 \times 10^{-7}$ | 0.581 | 0.565 |
| $D_s$ | -0.00314 | 0.00097 | -3.24 | 0.00243 |

Table XV: In the left top panel: best-estimate, standard error ( $\sigma$ ), t-statistic and $p$ -value for the parameters of the linear interpolation, for correlation of $\alpha$ with seasonal levels of vitamin D (variable name: $D_s$ ).
|  |  |
| --- | --- |
| $R^2$ | 0.212 |
| $N$ | 42 |
| Cross-correlation | -0.162 |

**Table XVI:**
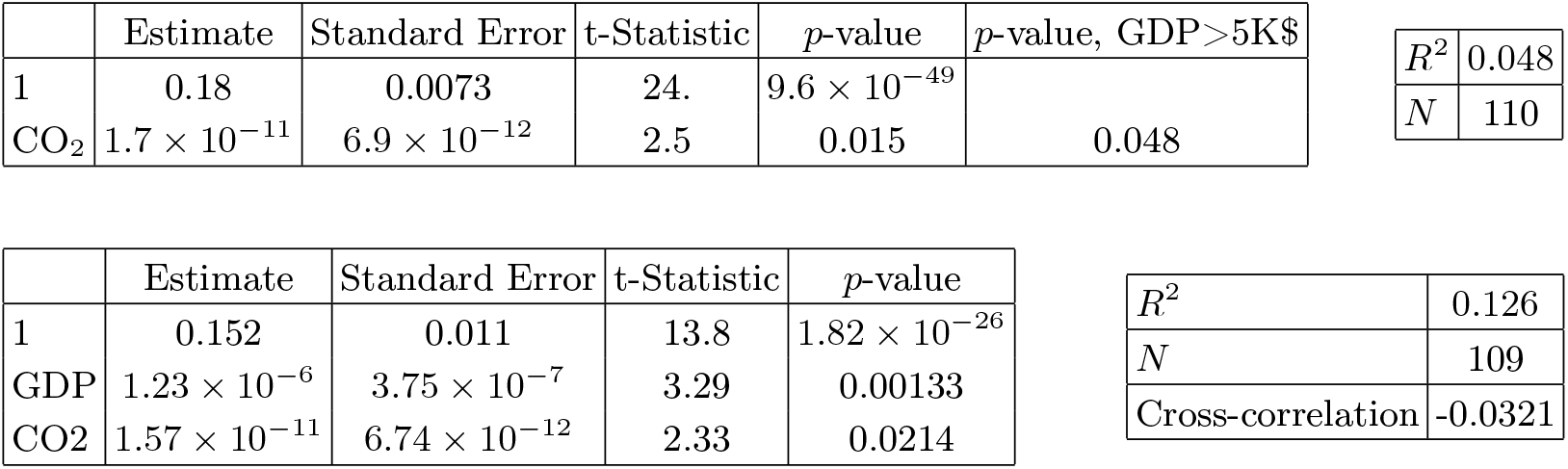
In the left top panel: best-estimate, standard error (*σ*), t-statistic and *p*-value for the parameters of the linear interpolation, for correlation of *α* with CO_2_ emissions. We also show the *p*-value, excluding countries below 5 thousand $ GDP per capita. In the left bottom panel: same quantities for correlation of *α* with CO_2_ and GDP per capita. In the right panels: *R*^2^ for the best-estimate and number of countries *N*. We also show the correlation coefficient between the 2 variables in the two-variable fit.

**Figure 16:**
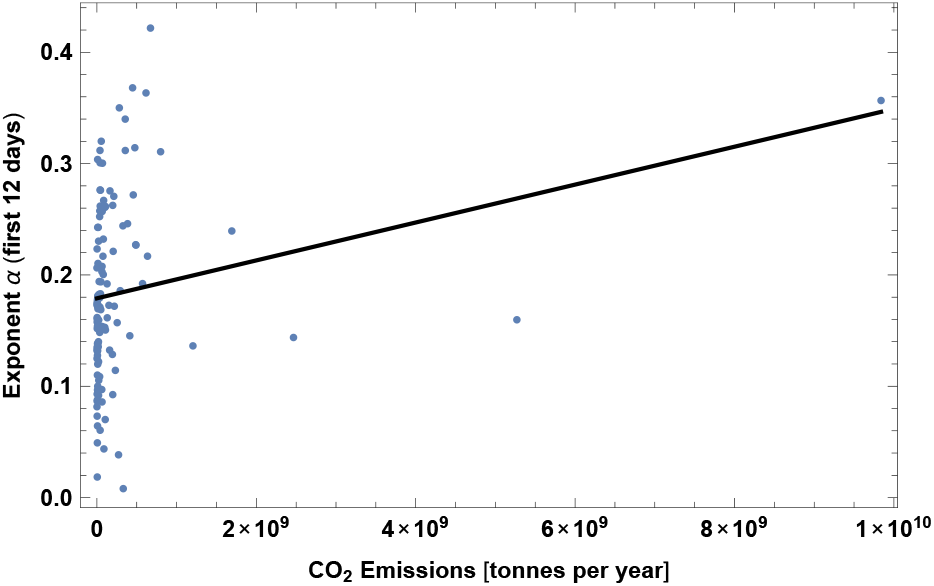
Exponent *α* for each country vs. CO_2_ emissions. We show the data points and the best-fit for the linear interpolation.

### 15. Prevalence of type-1 Diabetes

This is the prevalence of type-1 Diabetes in children, 0-19 years-old, taken from [23]. Results are shown in Fig. 17 and Table XXIII. Note however that significance becomes very small when restricting to countries with GDP per capita larger than 5K$. Note also that in the case of diabetes of *any* kind we do *not* find a correlation with COVID-19. Such correlation, even if not highly significant, could be non-trivial and could constitute useful information for clinical and genetic research. See also [49].

**Figure 17:**
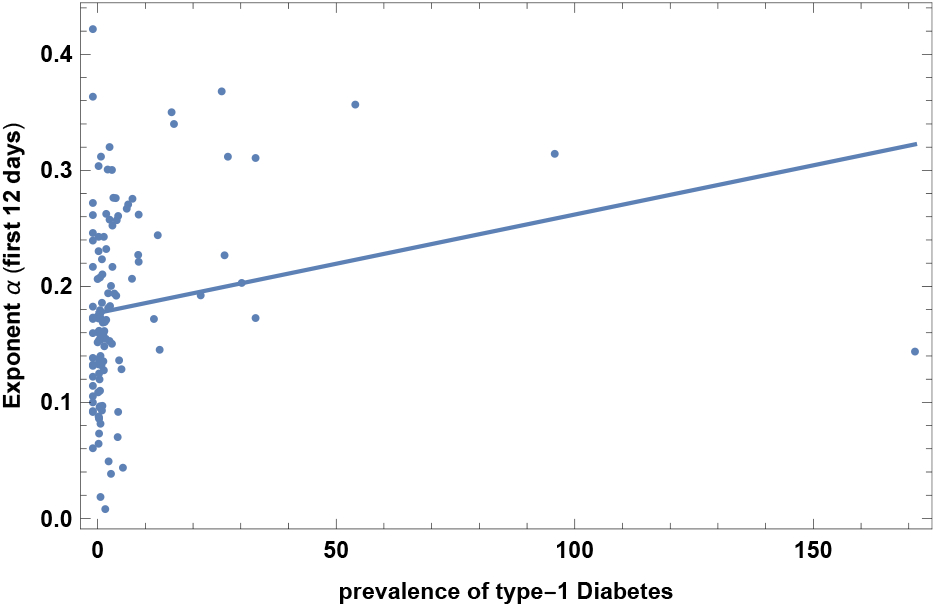
Exponent *α* for each country vs. prevalence of type-1 Diabetes. We show the data points and the best-fit for the linear interpolation.

### 16. Tuberculosis (BCG) vaccination coverage

This dataset is the vaccination coverage for tuberculosis for year 2015 [58]. Results are shown in Fig. 18 and Table XVIII. We find a negative correlation. Note that this depends mostly on the three countries with low coverage present in the plot: by excluding them the correlation becomes non-significant. Note also that several countries with no compulsory vaccination (such as USA or Italy) do not have an estimate for BCG coverage and were not included in the analysis. Including them, with very low coverage, would probably affect a lot the significance. This correlation, even if not highly significant and to be confirmed by more data, is also quite non-trivial and could be useful information for clinical and genetic research (see also [29–31]) and even for vaccine development [32–34].

**Table XVII:** In the left top panel: best-estimate, standard error (*σ*), t-statistic and *p*-value for the parameters of the linear interpolation, for correlation of *α* with prevalence of type-1 Diabetes, DIA. We also show the *p*-value, excluding countries below 5 thousand $ GDP per capita. In the left bottom panel: same quantities for correlation of *α* with DIA and GDP per capita. In the right panels: *R*^2^ for the best-estimate and number of countries *N*. We also show the correlation coefficient between the 2 variables in the two-variable fit.

| | Estimate | Standard Error | t-Statistic | $p$-value | $p$-value, GDP>5K\$ | $R^2$ | $N$ |
| --- | --- | --- | --- | --- | --- | --- | --- |
| 1 | 0.177 | 0.00793 | 22.4 | $2.35 \times 10^{-41}$ | | 0.052 | |
| DIAB | 0.00087 | 0.000367 | 2.37 | 0.0198 | 0.072 |  | 105 |

Table XVII: In the left top panel: best-estimate, standard error ( $\sigma$ ), t-statistic and $p$ -value for the parameters of the linear interpolation, for correlation of $\alpha$ with prevalence of type-1 Diabetes, DIA.
| | Estimate | Standard Error | t-Statistic | $p$-value | $R^2$ | $N$ |
| --- | --- | --- | --- | --- | --- | --- |
| 1 | 0.142 | 0.0116 | 12.3 | $1.52 \times 10^{-21}$ | 0.189 | |
| GDP | $1.58 \times 10^{-6}$ | $3.88 \times 10^{-7}$ | 4.08 | 0.000092 | | 101 |
| DIAB | 0.000932 | 0.000349 | 2.67 | 0.00889 | Cross-correlation | 0.0415 |

**Table XVIII:** In the left top panel: best-estimate, standard error (*σ*), t-statistic and *p*-value for the parameters of the linear interpolation, for correlation of *α* with BCG vaccination coverage (BCG). We also show the *p*-value, excluding countries below 5 thousand $ GDP per capita. In the left bottom panel: same quantities for correlation of *α* with BCG and GDP per capita. In the right panels: *R*^2^ for the best-estimate and number of countries *N*. We also show the correlation coefficient between the 2 variables in the two-variable fit.

| | Estimate | Standard Error | t-Statistic | $p$-value | $p$-value, GDP>5K\$ | $R^2$ | $N$ |
| --- | --- | --- | --- | --- | --- | --- | --- |
| 1 | 0.289 | 0.053 | 5.46 | $3.99 \times 10^{-7}$ | | 0.051 | |
| BCG | -0.00129 | 0.000579 | -2.22 | 0.0286 | 0.011 |  | 94 |

Table XVIII: In the left top panel: best-estimate, standard error ( $\sigma$ ), t-statistic and $p$ -value for the parameters of the linear interpolation, for correlation of $\alpha$ with BCG vaccination coverage (BCG).
| | Estimate | Standard Error | t-Statistic | $p$-value | $R^2$ | $N$ |
| --- | --- | --- | --- | --- | --- | --- |
| 1 | 0.28 | 0.0534 | 5.24 | $1.07 \times 10^{-6}$ | 0.084 | |
| GDP | $8.53 \times 10^{-7}$ | $4.79 \times 10^{-7}$ | 1.78 | 0.0781 | | 92 |
| BCG | -0.00134 | 0.00058 | -2.3 | 0.0235 | Cross-correlation | -0.0367 |

**Figure 18:**
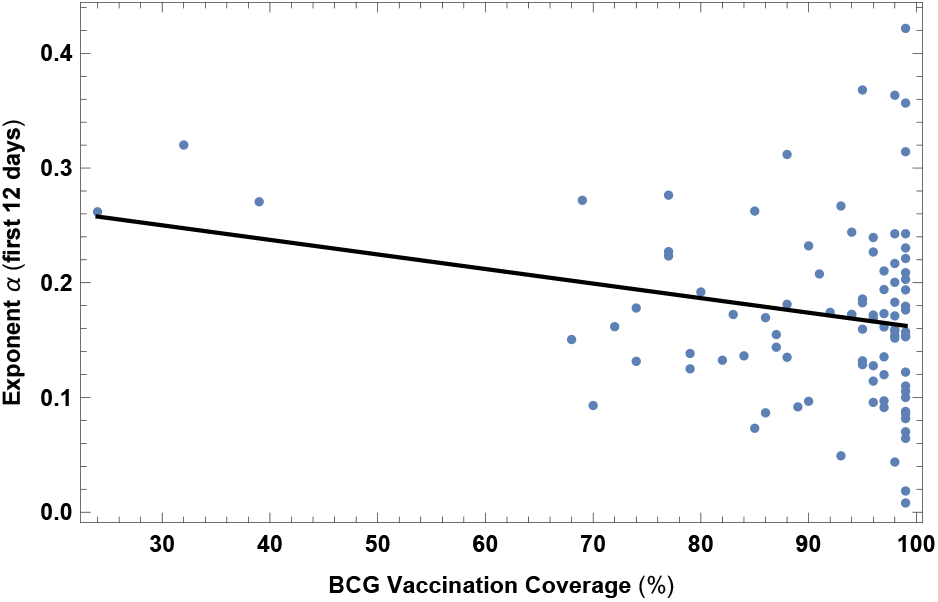
Exponent *α* for each country vs. BCG vaccination coverage. We show the data points and the best-fit for the linear interpolation.

#### A. “Counterintuitive” correlations

We show here other correlations that are somehow counterintuitive, since they go in the opposite direction than from a naive expectation. We will try to interpret these results in section V.

### 1. Death rate from air pollution

This dataset is for year 2015. Results are shown in Fig. 19 and Table XIX. Contrary to naive expectations and to claims in the opposite direction [48], we find that countries with larger death rate from air pollution actually have *slower* COVID-19 contagion.

**Figure 19:**
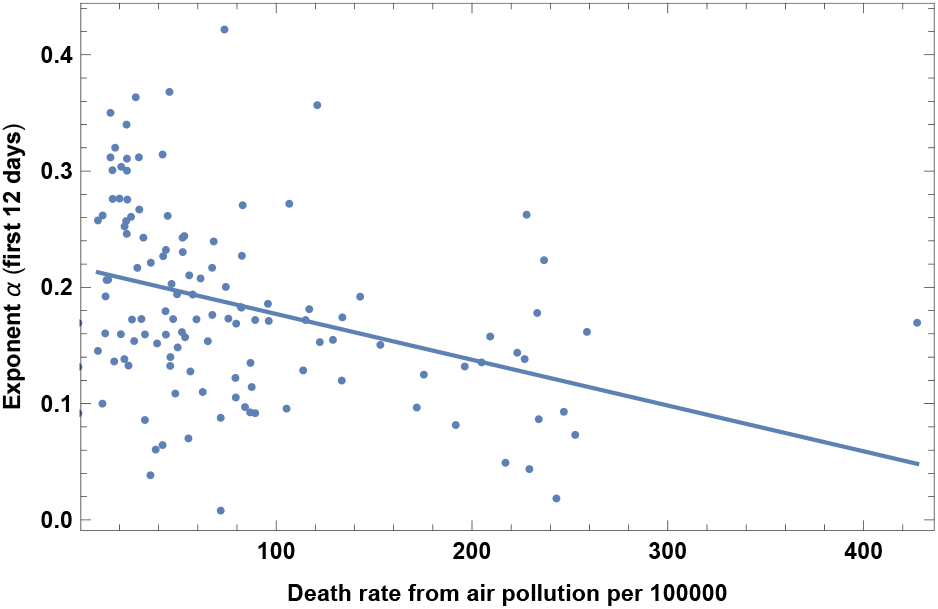
Exponent *α* for each country vs. death rate from air pollution per 100000. We show the data points and the best-fit for the linear interpolation.

**Table XIX:** In the left top panel: best-estimate, standard error (*σ*), t-statistic and *p*-value for the parameters of the linear interpolation, for correlation of *α* with death rate from air pollution per 100000 (POLL). We also show the *p*-value, excluding countries below 5 thousand $ GDP per capita. In the left bottom panel: same quantities for correlation of *α* with POLL and GDP per capita. In the right panels: *R*^2^ for the best-estimate and number of countries *N*. We also show the correlation coefficient between the 2 variables in the two-variable fit.

| | Estimate | Standard Error | t-Statistic | $p$-value | $p$-value, GDP>5K\$ | | |
| --- | --- | --- | --- | --- | --- | --- | --- |
| 1 | 0.216 | 0.0102 | 21.3 | $8.97 \times 10^{-43}$ | | $R^2$ | 0.132 |
| POLL | -0.000394 | 0.0000917 | -4.29 | 0.0000357 | 0.040 | $N$ | 123 |

Table XIX: In the left top panel: best-estimate, standard error ( $\sigma$ ), t-statistic and $p$ -value for the parameters of the linear interpolation, for correlation of $\alpha$ with death rate from air pollution per 100000 (POLL).
| | Estimate | Standard Error | t-Statistic | $p$-value | | |
| --- | --- | --- | --- | --- | --- | --- |
| 1 | 0.211 | 0.0204 | 10.3 | $3.75 \times 10^{-18}$ | $R^2$ | 0.158 |
| GDP | $3.13 \times 10^{-7}$ | $4.77 \times 10^{-7}$ | 0.655 | 0.514 | $N$ | 120 |
| POLL | -0.000418 | 0.000131 | -3.19 | 0.00185 | Cross-correlation | 0.630 |

### 2. High blood pressure in females

This dataset is for year 2015. Results are shown in Fig. 20 and Table XX. Countries with larger share of high blood pressure in females have *slower* COVID-19 contagion. Note that we do *not* find a significant correlation instead with high blood pressure in *males*.

**Figure 20:**
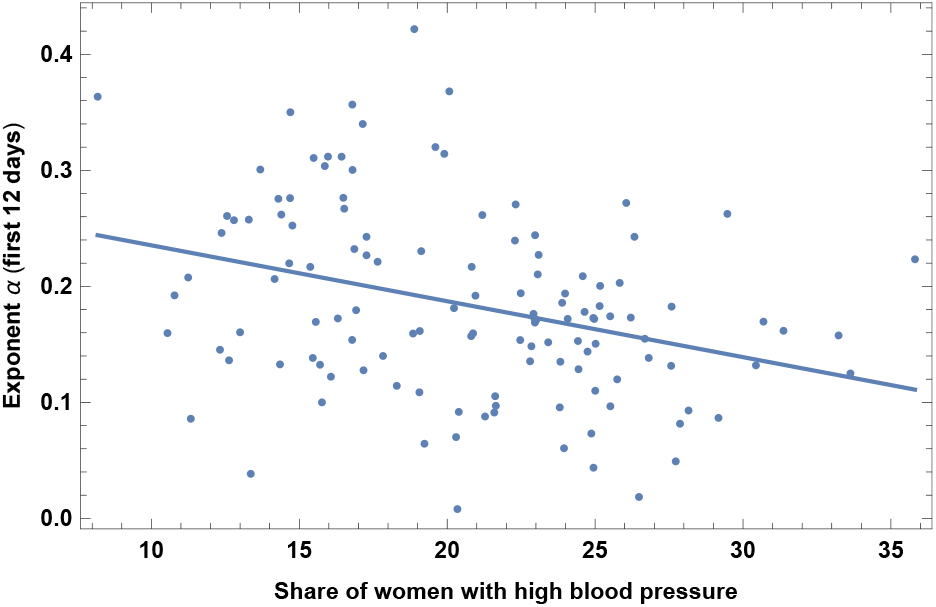
Exponent *α* for each country vs. share of women with high blood pressure. We show the data points and the best-fit for the linear interpolation.

**Table XX:**
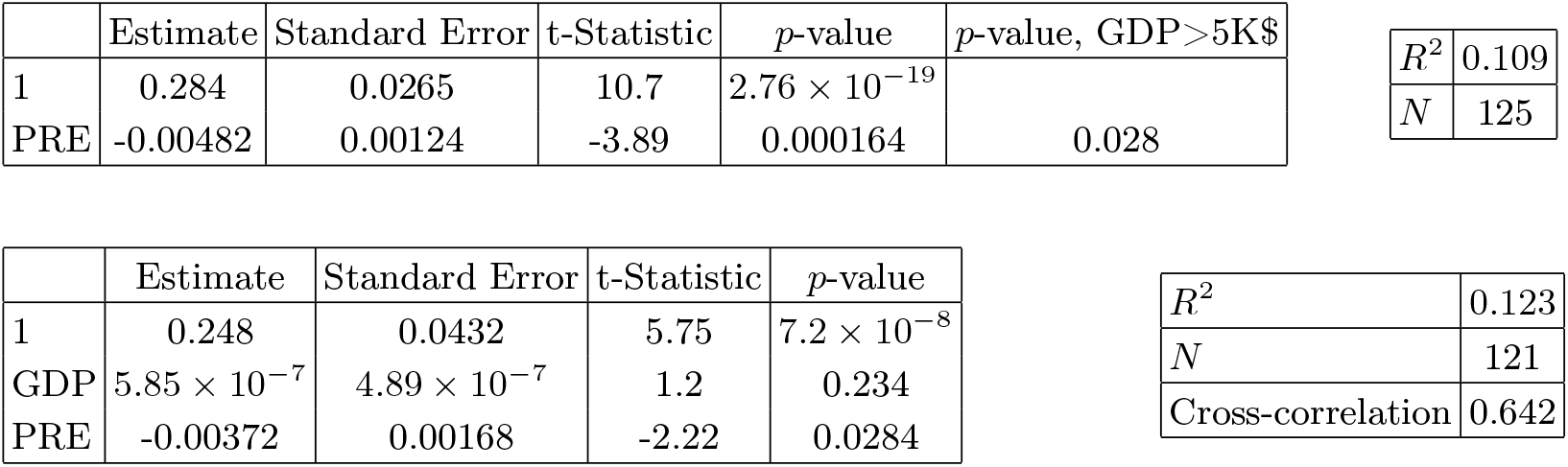
In the left top panel: best-estimate, standard error (*σ*), t-statistic and *p*-value for the parameters of the linear interpolation, for correlation of *α* with share of women with high blood pressure (PRE). We also show the *p*-value, excluding countries below 5 thousand $ GDP per capita. In the left bottom panel: same quantities for correlation of *α* with PRE and GDP per capita. In the right panels: *R*^2^ for the best-estimate and number of countries *N*. We also show the correlation coefficient between the 2 variables in the two-variable fit.

| | Estimate | Standard Error | t-Statistic | $p$-value | $p$-value, GDP > 5K\$ |
| --- | --- | --- | --- | --- | --- |
| 1 | 0.284 | 0.0265 | 10.7 | $2.76 \times 10^{-19}$ | |
| PRE | -0.00482 | 0.00124 | -3.89 | 0.000164 | 0.028 |

|  |  |
| --- | --- |
| $R^2$ | 0.109 |
| $N$ | 125 |

| | Estimate | Standard Error | t-Statistic | $p$-value |
| --- | --- | --- | --- | --- |
| 1 | 0.248 | 0.0432 | 5.75 | $7.2 \times 10^{-8}$ |
| GDP | $5.85 \times 10^{-7}$ | $4.89 \times 10^{-7}$ | 1.2 | 0.234 |
| PRE | -0.00372 | 0.00168 | -2.22 | 0.0284 |

Table XX: In the left top panel: best-estimate, standard error ( $\sigma$ ), t-statistic and $p$ -value for the parameters of the linear interpolation, for correlation of $\alpha$ with share of women with high blood pressure (PRE).
|  |  |
| --- | --- |
| $R^2$ | 0.123 |
| $N$ | 121 |
| Cross-correlation | 0.642 |

### 3. Hepatitis B incidence rate

This is the incidence of hepatitis B, measured as the number of new cases of hepatitis B per 100,000 individuals in a given population, for the year 2015. Results are shown in Fig. 21 and Table XXI. Countries with higher incidence of hepatitis B have *slower* contagion of COVID-19.

**Figure 21:**
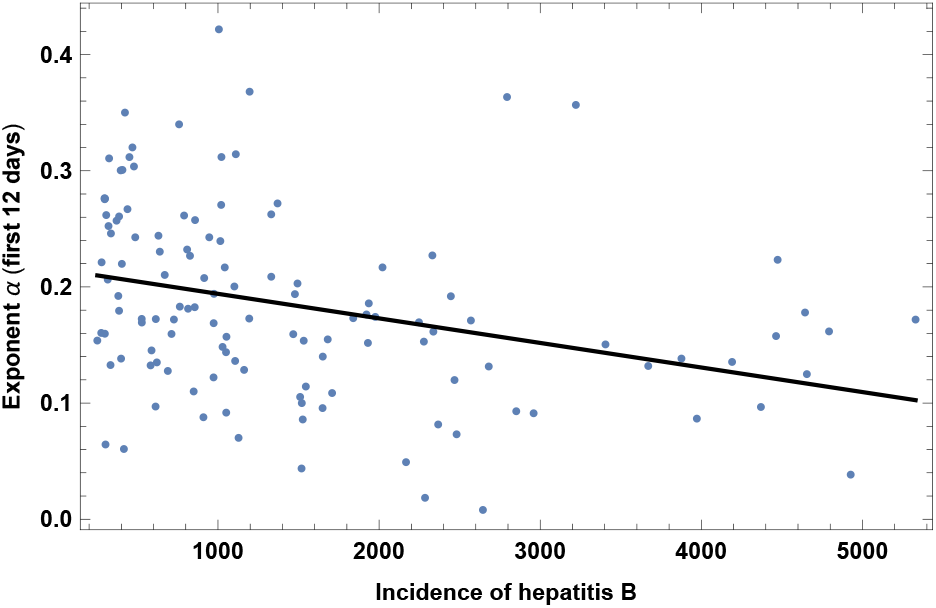
Exponent *α* for each country vs. incidence of Hepatitis B, for the relevant period of time, as defined in the text, for the base set of 42 countries. We show the data points and the best-fit for the linear interpolation.

**Table XXI:**
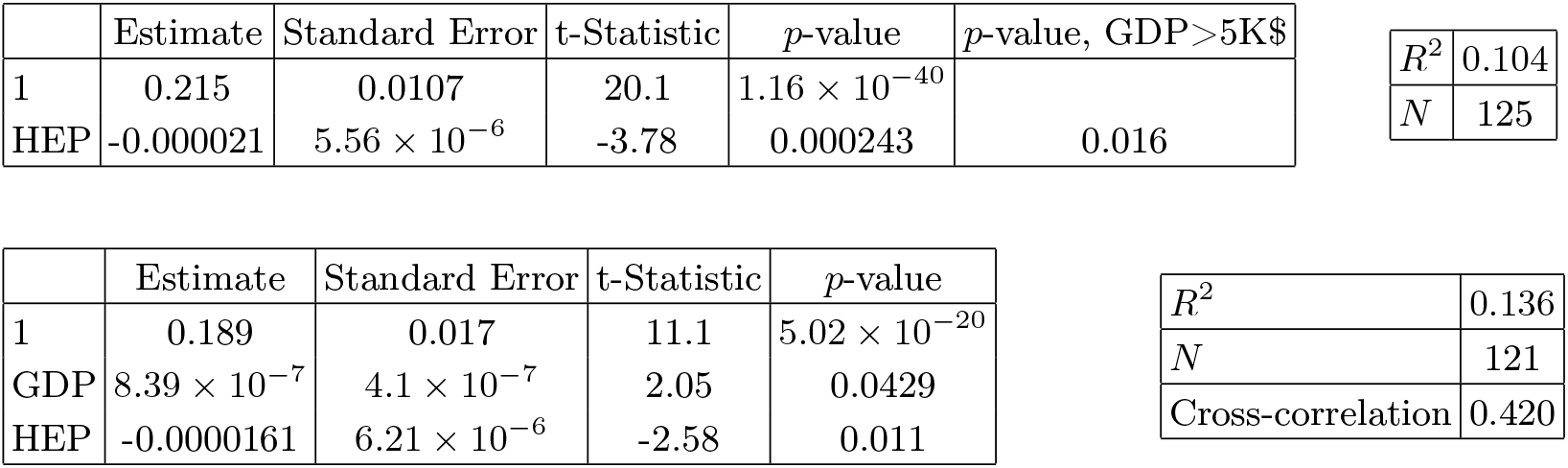
In the left top panel: best-estimate, standard error (*σ*), t-statistic and *p*-value for the parameters of the linear interpolation, for correlation of *α* with incidence of Hepatitis B (HEP). We also show the *p*-value, excluding countries below 5 thousand $ GDP per capita. In the left bottom panel: same quantities for correlation of *α* with HEP and GDP per capita. In the right panels: *R*^2^ for the best-estimate and number of countries *N*. We also show the correlation coefficient between the 2 variables in the two-variable fit.

| | Estimate | Standard Error | t-Statistic | $p$-value | $p$-value, GDP>5K\$ |
| --- | --- | --- | --- | --- | --- |
| 1 | 0.215 | 0.0107 | 20.1 | $1.16 \times 10^{-40}$ | |
| HEP | -0.000021 | $5.56 \times 10^{-6}$ | -3.78 | 0.000243 | 0.016 |

|  |  |
| --- | --- |
| $R^2$ | 0.104 |
| $N$ | 125 |

| | Estimate | Standard Error | t-Statistic | $p$-value |
| --- | --- | --- | --- | --- |
| 1 | 0.189 | 0.017 | 11.1 | $5.02 \times 10^{-20}$ |
| GDP | $8.39 \times 10^{-7}$ | $4.1 \times 10^{-7}$ | 2.05 | 0.0429 |
| HEP | -0.0000161 | $6.21 \times 10^{-6}$ | -2.58 | 0.011 |

Table XXI: In the left top panel: best-estimate, standard error ( $\sigma$ ), t-statistic and $p$ -value for the parameters of the linear interpolation, for correlation of $\alpha$ with incidence of Hepatitis B (HEP).
|  |  |
| --- | --- |
| $R^2$ | 0.136 |
| $N$ | 121 |
| Cross-correlation | 0.420 |

### 4. Prevalence of Anemia

Prevalence of anemia in children in 2016, measured as the share of children under the age of five with hemoglobin levels less than 110 grams per liter at sea level. A similar but less significant correlation is found also with anemia in adults, which we do not report here. Results are shown in Fig. 22 and Table XXII. The significance is quite high, but could be interpreted as due to a high correlation with life expectancy, as we explain in section V. A different hypothesis is that this might be related to genetic factors, which might affect the immune response to COVID-19.

**Figure 22:**
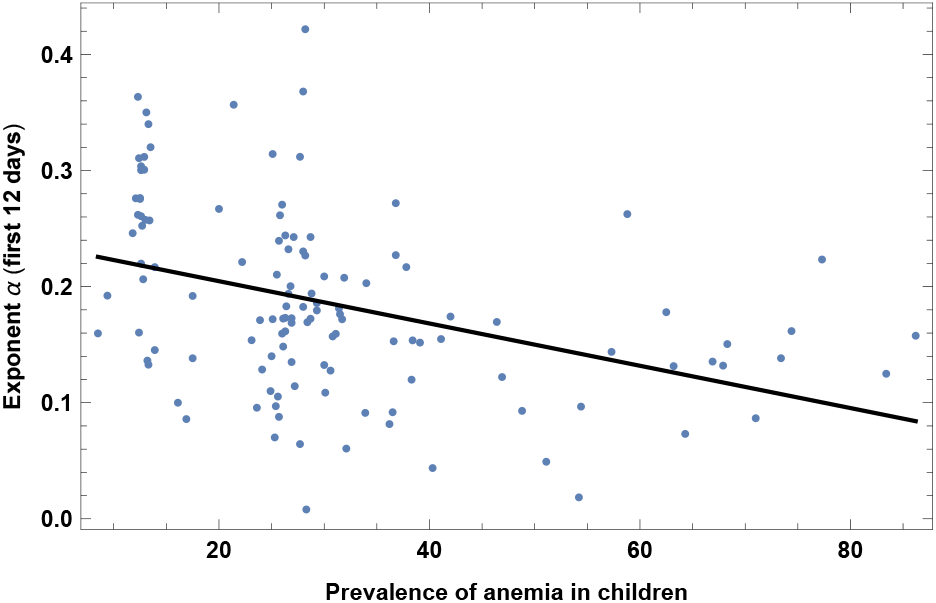
Exponent *α* for each country vs. prevalence of anemia in children. We show the data points and the best-fit for the linear interpolation.

**Table XXII:**
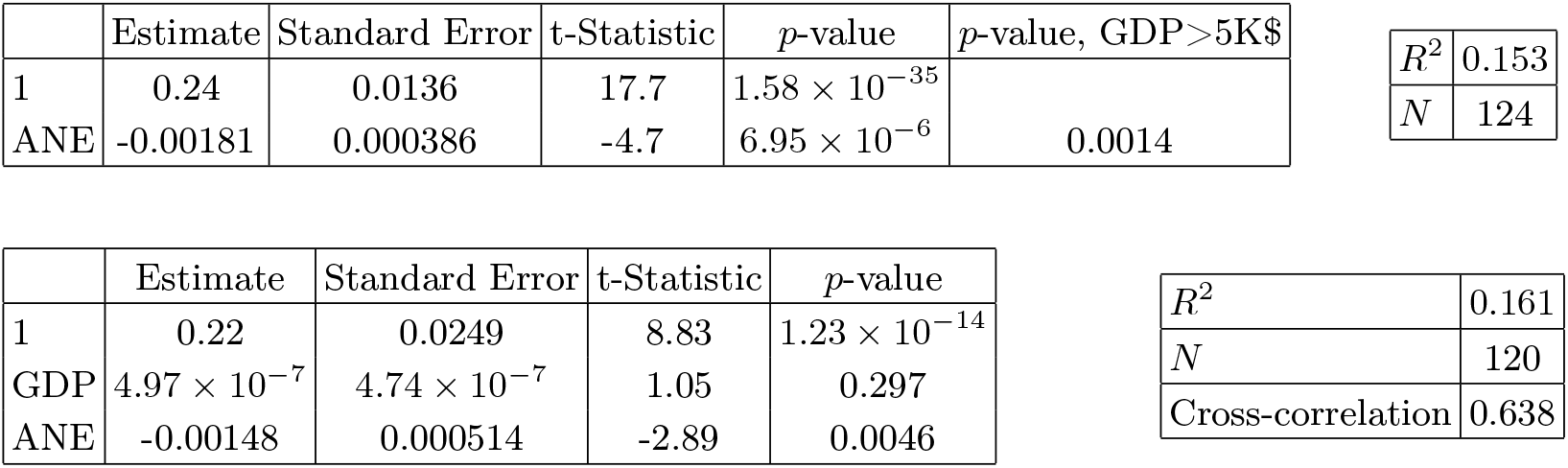
In the left top panel: best-estimate, standard error (*σ*), t-statistic and *p*-value for the parameters of the linear interpolation, for correlation of *α* with prevalence of anemia in children (ANE). We also show the *p*-value, excluding countries below 5 thousand $ GDP per capita. In the left bottom panel: same quantities for correlation of *α* with ANE and GDP per capita. In the right panels: *R*^2^ for the best-estimate and number of countries *N*. We also show the correlation coefficient between the 2 variables in the two-variable fit.

| | Estimate | Standard Error | t-Statistic | $p$-value | $p$-value, GDP>5K\$ |
| --- | --- | --- | --- | --- | --- |
| 1 | 0.24 | 0.0136 | 17.7 | $1.58 \times 10^{-35}$ | |
| ANE | -0.00181 | 0.000386 | -4.7 | $6.95 \times 10^{-6}$ | 0.0014 |

|  |  |
| --- | --- |
| $R^2$ | 0.153 |
| $N$ | 124 |

| | Estimate | Standard Error | t-Statistic | $p$-value |
| --- | --- | --- | --- | --- |
| 1 | 0.22 | 0.0249 | 8.83 | $1.23 \times 10^{-14}$ |
| GDP | $4.97 \times 10^{-7}$ | $4.74 \times 10^{-7}$ | 1.05 | 0.297 |
| ANE | -0.00148 | 0.000514 | -2.89 | 0.0046 |

Table XXII: In the left top panel: best-estimate, standard error ( $\sigma$ ), t-statistic and $p$ -value for the parameters of the linear interpolation, for correlation of $\alpha$ with prevalence of anemia in children (ANE).
|  |  |
| --- | --- |
| $R^2$ | 0.161 |
| $N$ | 120 |
| Cross-correlation | 0.638 |

**Table XXIII:**
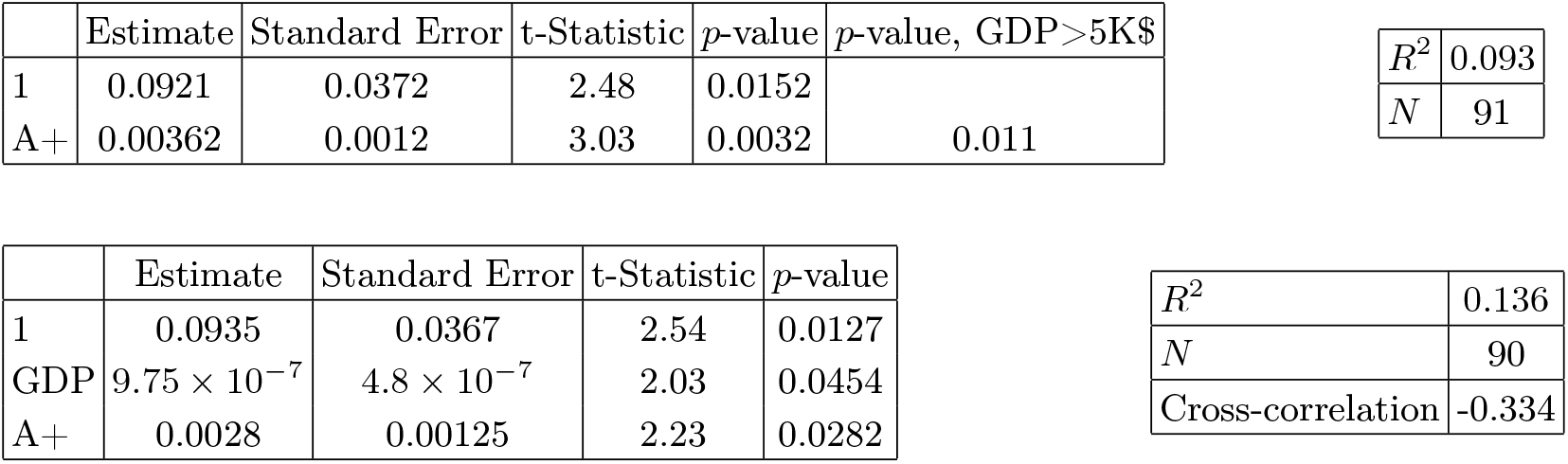
In the left top panel: best-estimate, standard error (*σ*), t-statistic and *p*-value for the parameters of the linear interpolation, for correlation of *α* with percentage of population with blood type A+. We also show the *p*-value, excluding countries below 5 thousand $ GDP per capita. In the left bottom panel: same quantities for correlation of *α* with A+ blood and GDP per capita. In the right panels: *R*^2^ for the best-estimate and number of countries *N*. We also show the correlation coefficient between the 2 variables in the two-variable fit.

| | Estimate | Standard Error | t-Statistic | $p$-value | $p$-value, GDP>5K\$ |
| --- | --- | --- | --- | --- | --- |
| 1 | 0.0921 | 0.0372 | 2.48 | 0.0152 |  |
| A+ | 0.00362 | 0.0012 | 3.03 | 0.0032 | 0.011 |

|  |  |
| --- | --- |
| $R^2$ | 0.093 |
| $N$ | 91 |

| | Estimate | Standard Error | t-Statistic | $p$-value |
| --- | --- | --- | --- | --- |
| 1 | 0.0935 | 0.0367 | 2.54 | 0.0127 |
| GDP | $9.75 \times 10^{-7}$ | $4.8 \times 10^{-7}$ | 2.03 | 0.0454 |
| A+ | 0.0028 | 0.00125 | 2.23 | 0.0282 |

Table XXIII: In the left top panel: best-estimate, standard error ( $\sigma$ ), t-statistic and $p$ -value for the parameters of the linear interpolation, for correlation of $\alpha$ with percentage of population with blood type A+.
|  |  |
| --- | --- |
| $R^2$ | 0.136 |
| $N$ | 90 |
| Cross-correlation | -0.334 |

**Table XXIV:**
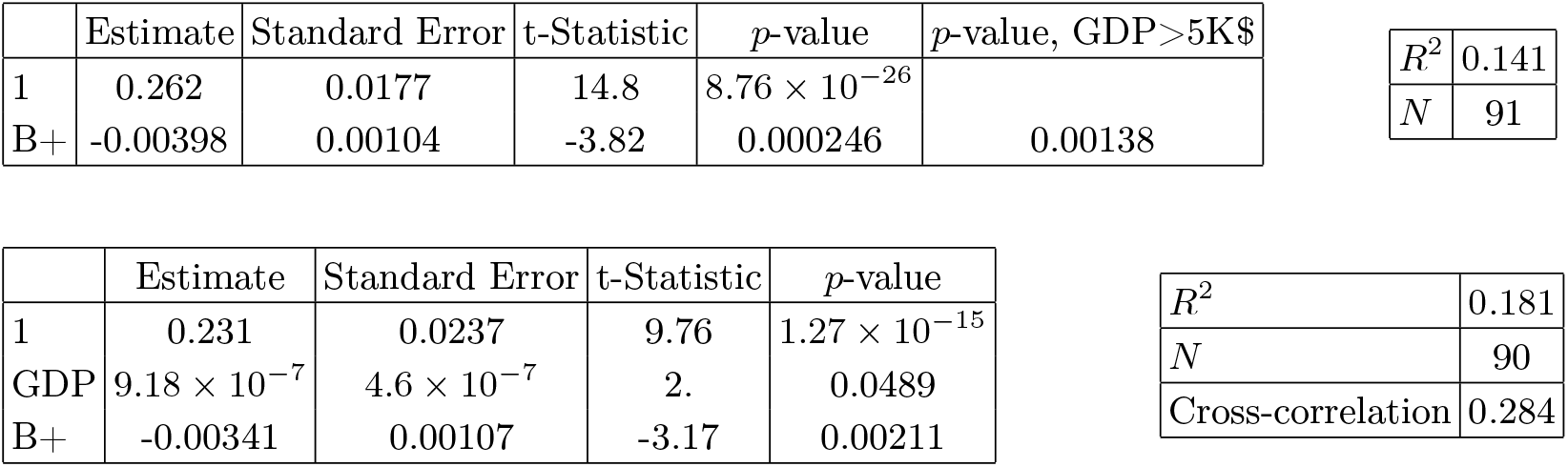
In the left top panel: best-estimate, standard error (*σ*), t-statistic and *p*-value for the parameters of the linear interpolation, for correlation of *α* with percentage of population with blood type B+. We also show the *p*-value, excluding countries below 5 thousand $ GDP per capita. In the left bottom panel: same quantities for correlation of *α* with B+ and GDP per capita. In the right panels: *R*^2^ for the best-estimate and number of countries *N*. We also show the correlation coefficient between the 2 variables in the two-variable fit.

| | Estimate | Standard Error | t-Statistic | $p$-value | $p$-value, GDP>5K\$ |
| --- | --- | --- | --- | --- | --- |
| 1 | 0.262 | 0.0177 | 14.8 | $8.76 \times 10^{-26}$ | |
| B+ | -0.00398 | 0.00104 | -3.82 | 0.000246 | 0.00138 |

|  |  |
| --- | --- |
| $R^2$ | 0.141 |
| $N$ | 91 |

| | Estimate | Standard Error | t-Statistic | $p$-value |
| --- | --- | --- | --- | --- |
| 1 | 0.231 | 0.0237 | 9.76 | $1.27 \times 10^{-15}$ |
| GDP | $9.18 \times 10^{-7}$ | $4.6 \times 10^{-7}$ | 2. | 0.0489 |
| B+ | -0.00341 | 0.00107 | -3.17 | 0.00211 |

Table XXIV: In the left top panel: best-estimate, standard error ( $\sigma$ ), t-statistic and $p$ -value for the parameters of the linear interpolation, for correlation of $\alpha$ with percentage of population with blood type B+.
|  |  |
| --- | --- |
| $R^2$ | 0.181 |
| $N$ | 90 |
| Cross-correlation | 0.284 |

**Table XXV:**
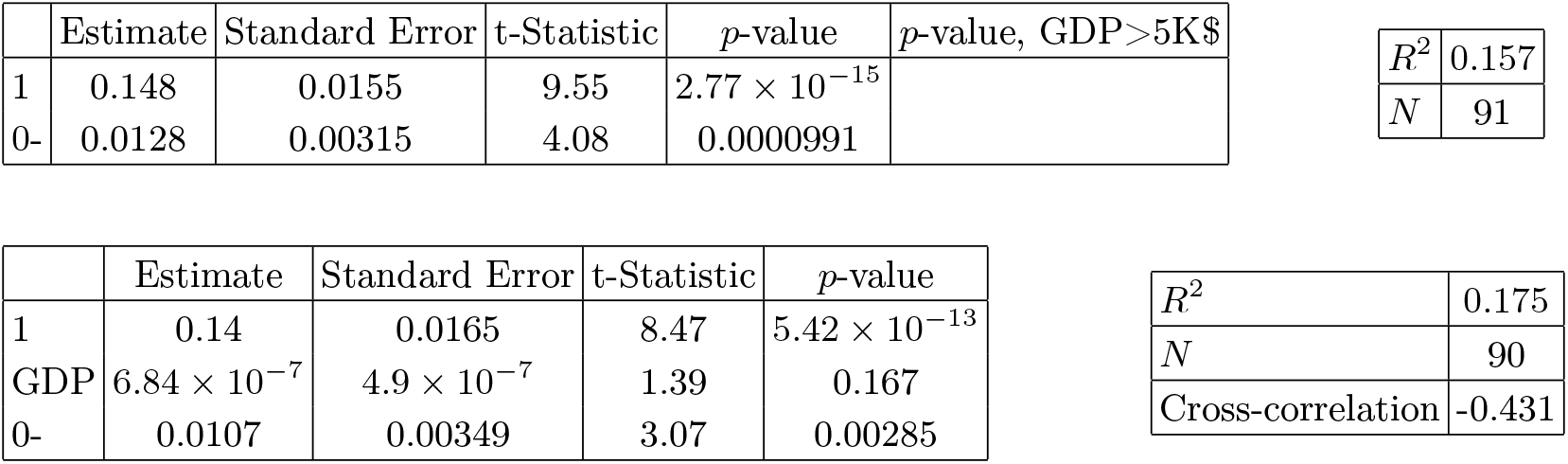
In the left top panel: best-estimate, standard error (*σ*), t-statistic and *p*-value for the parameters of the linear interpolation, for correlation of *α* with percentage of population with blood type 0−. We also show the *p*-value, excluding countries below 5 thousand $ GDP per capita. In the left bottom panel: same quantities for correlation of *α* with 0−and GDP per capita. In the right panels: *R*^2^ for the best-estimate and number of countries *N*. We also show the correlation coefficient between the 2 variables in the two-variable fit.

| | Estimate | Standard Error | t-Statistic | $p$-value | $p$-value, GDP>5K\$ |
| --- | --- | --- | --- | --- | --- |
| 1 | 0.148 | 0.0155 | 9.55 | $2.77 \times 10^{-15}$ | |
| 0- | 0.0128 | 0.00315 | 4.08 | 0.0000991 |  |

|  |  |
| --- | --- |
| $R^2$ | 0.157 |
| $N$ | 91 |

| | Estimate | Standard Error | t-Statistic | $p$-value |
| --- | --- | --- | --- | --- |
| 1 | 0.14 | 0.0165 | 8.47 | $5.42 \times 10^{-13}$ |
| GDP | $6.84 \times 10^{-7}$ | $4.9 \times 10^{-7}$ | 1.39 | 0.167 |
| 0- | 0.0107 | 0.00349 | 3.07 | 0.00285 |

Table XXV: In the left top panel: best-estimate, standard error ( $\sigma$ ), t-statistic and $p$ -value for the parameters of the linear interpolation, for correlation of $\alpha$ with percentage of population with blood type 0-.
|  |  |
| --- | --- |
| $R^2$ | 0.175 |
| $N$ | 90 |
| Cross-correlation | -0.431 |

**Table XXVI:**
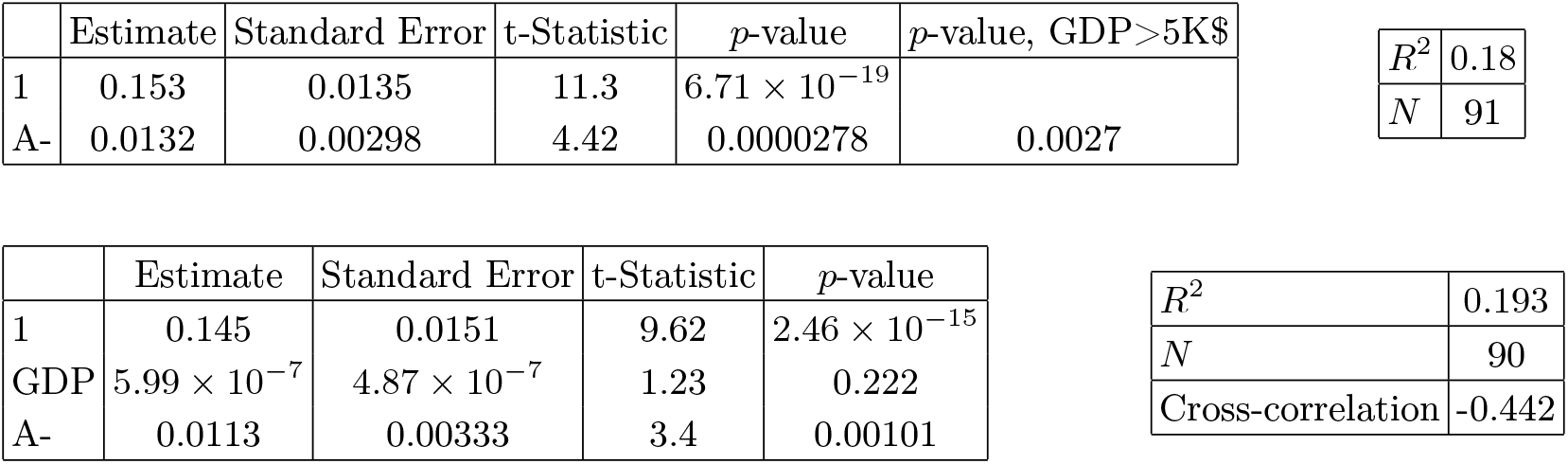
In the left top panel: best-estimate, standard error (*σ*), t-statistic and *p*-value for the parameters of the linear interpolation, for correlation of *α* with percentage of population with blood type A-. We also show the *p*-value, excluding countries below 5 thousand $ GDP per capita. In the left bottom panel: same quantities for correlation of *α* with A-and GDP per capita. In the right panels: *R*^2^ for the best-estimate and number of countries *N*. We also show the correlation coefficient between the 2 variables in the two-variable fit.

| | Estimate | Standard Error | t-Statistic | $p$-value | $p$-value, GDP>5K\$ |
| --- | --- | --- | --- | --- | --- |
| 1 | 0.153 | 0.0135 | 11.3 | $6.71 \times 10^{-19}$ | |
| A- | 0.0132 | 0.00298 | 4.42 | 0.0000278 | 0.0027 |

|  |  |
| --- | --- |
| $R^2$ | 0.18 |
| $N$ | 91 |

| | Estimate | Standard Error | t-Statistic | $p$-value |
| --- | --- | --- | --- | --- |
| 1 | 0.145 | 0.0151 | 9.62 | $2.46 \times 10^{-15}$ |
| GDP | $5.99 \times 10^{-7}$ | $4.87 \times 10^{-7}$ | 1.23 | 0.222 |
| A- | 0.0113 | 0.00333 | 3.4 | 0.00101 |

Table XXVI: In the left top panel: best-estimate, standard error ( $\sigma$ ), t-statistic and $p$ -value for the parameters of the linear interpolation, for correlation of $\alpha$ with percentage of population with blood type A-.
|  |  |
| --- | --- |
| $R^2$ | 0.193 |
| $N$ | 90 |
| Cross-correlation | -0.442 |

**Table XXVII:**
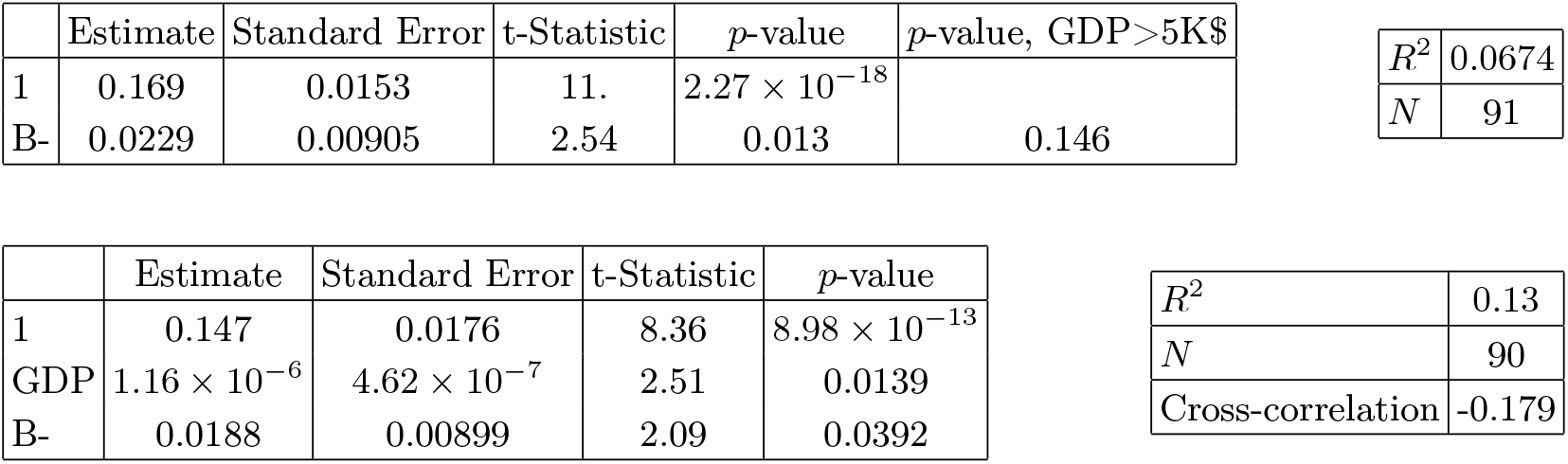
In the left top panel: best-estimate, standard error (*σ*), t-statistic and *p*-value for the parameters of the linear interpolation, for correlation of *α* with percentage of population with blood type B-. We also show the *p*-value, excluding countries below 5 thousand $ GDP per capita. In the left bottom panel: same quantities for correlation of *α* with B- and GDP per capita. In the right panels: *R*^2^ for the best-estimate and number of countries *N*. We also show the correlation coefficient between the 2 variables in the two-variable fit.

| | Estimate | Standard Error | t-Statistic | $p$-value | $p$-value, GDP>5K\$ |
| --- | --- | --- | --- | --- | --- |
| 1 | 0.169 | 0.0153 | 11. | $2.27 \times 10^{-18}$ | |
| B- | 0.0229 | 0.00905 | 2.54 | 0.013 | 0.146 |

|  |  |
| --- | --- |
| $R^2$ | 0.0674 |
| $N$ | 91 |

| | Estimate | Standard Error | t-Statistic | $p$-value |
| --- | --- | --- | --- | --- |
| 1 | 0.147 | 0.0176 | 8.36 | $8.98 \times 10^{-13}$ |
| GDP | $1.16 \times 10^{-6}$ | $4.62 \times 10^{-7}$ | 2.51 | 0.0139 |
| B- | 0.0188 | 0.00899 | 2.09 | 0.0392 |

Table XXVII: In the left top panel: best-estimate, standard error ( $\sigma$ ), t-statistic and $p$ -value for the parameters of the linear interpolation, for correlation of $\alpha$ with percentage of population with blood type B-.
|  |  |
| --- | --- |
| $R^2$ | 0.13 |
| $N$ | 90 |
| Cross-correlation | -0.179 |

**Table XXVIII:**
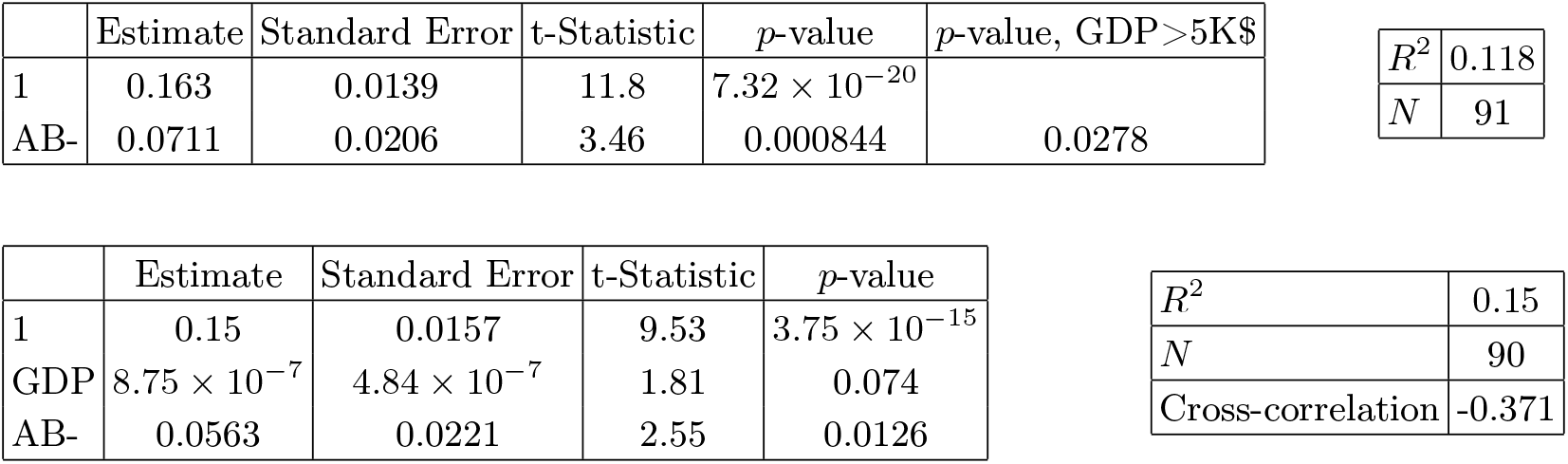
In the left top panel: best-estimate, standard error (*σ*), t-statistic and *p*-value for the parameters of the linear interpolation, for correlation of *α* with percentage of population with blood type AB-. We also show the *p*-value, excluding countries below 5 thousand $ GDP per capita. In the left bottom panel: same quantities for correlation of *α* with AB- and GDP per capita. In the right panels: *R*^2^ for the best-estimate and number of countries *N*. We also show the correlation coefficient between the 2 variables in the two-variable fit.

| | Estimate | Standard Error | t-Statistic | $p$-value | $p$-value, GDP>5K\$ |
| --- | --- | --- | --- | --- | --- |
| 1 | 0.163 | 0.0139 | 11.8 | $7.32 \times 10^{-20}$ | |
| AB- | 0.0711 | 0.0206 | 3.46 | 0.000844 | 0.0278 |

|  |  |
| --- | --- |
| $R^2$ | 0.118 |
| $N$ | 91 |

| | Estimate | Standard Error | t-Statistic | $p$-value |
| --- | --- | --- | --- | --- |
| 1 | 0.15 | 0.0157 | 9.53 | $3.75 \times 10^{-15}$ |
| GDP | $8.75 \times 10^{-7}$ | $4.84 \times 10^{-7}$ | 1.81 | 0.074 |
| AB- | 0.0563 | 0.0221 | 2.55 | 0.0126 |

Table XXVIII: In the left top panel: best-estimate, standard error ( $\sigma$ ), t-statistic and $p$ -value for the parameters of the linear interpolation, for correlation of $\alpha$ with percentage of population with blood type AB-.
|  |  |
| --- | --- |
| $R^2$ | 0.15 |
| $N$ | 90 |
| Cross-correlation | -0.371 |

**Table XXIX:**
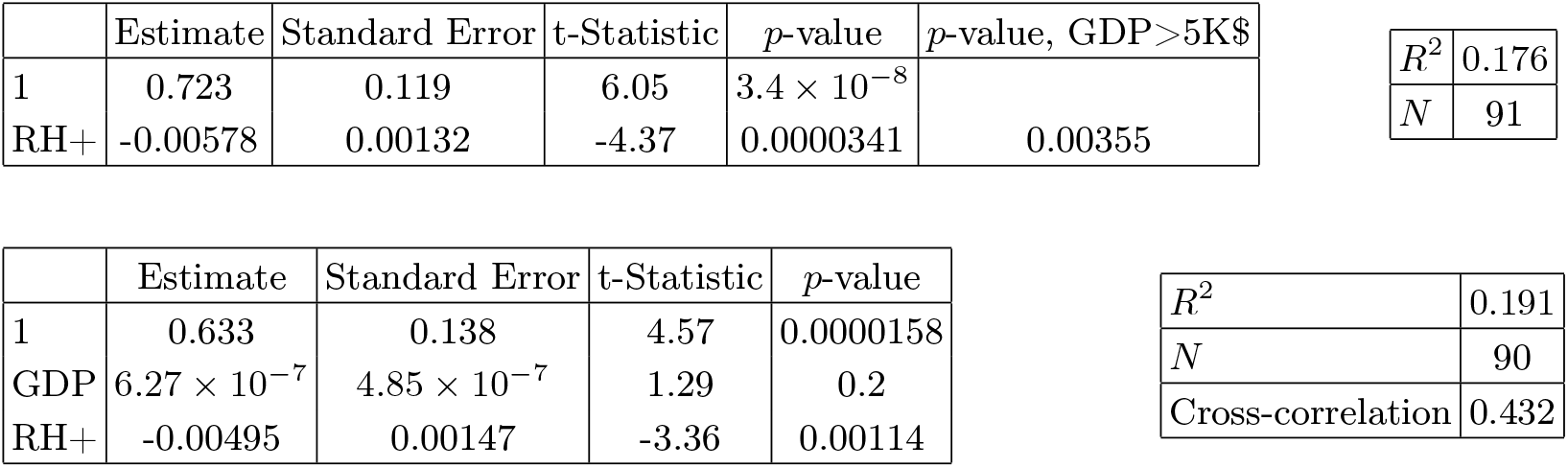
In the left top panel: best-estimate, standard error (*σ*), t-statistic and *p*-value for the parameters of the linear interpolation, for correlation of *α* with percentage of population with RH-positive blood (RH+). We also show the *p*-value, excluding countries below 5 thousand $ GDP per capita. In the left bottom panel: same quantities for correlation of *α* with RH+ and GDP per capita. In the right panels: *R*^2^ for the best-estimate and number of countries *N*. We also show the correlation coefficient between the 2 variables in the two-variable fit.

| | Estimate | Standard Error | t-Statistic | $p$-value | $p$-value, GDP>5K\$ |
| --- | --- | --- | --- | --- | --- |
| 1 | 0.723 | 0.119 | 6.05 | $3.4 \times 10^{-8}$ | |
| RH+ | -0.00578 | 0.00132 | -4.37 | 0.0000341 | 0.00355 |

|  |  |
| --- | --- |
| $R^2$ | 0.176 |
| $N$ | 91 |

| | Estimate | Standard Error | t-Statistic | $p$-value |
| --- | --- | --- | --- | --- |
| 1 | 0.633 | 0.138 | 4.57 | 0.0000158 |
| GDP | $6.27 \times 10^{-7}$ | $4.85 \times 10^{-7}$ | 1.29 | 0.2 |
| RH+ | -0.00495 | 0.00147 | -3.36 | 0.00114 |

Table XXIX: In the left top panel: best-estimate, standard error ( $\sigma$ ), t-statistic and $p$ -value for the parameters of the linear interpolation, for correlation of $\alpha$ with percentage of population with RH-positive blood (RH+).
|  |  |
| --- | --- |
| $R^2$ | 0.191 |
| $N$ | 90 |
| Cross-correlation | 0.432 |

**Table XXX:**
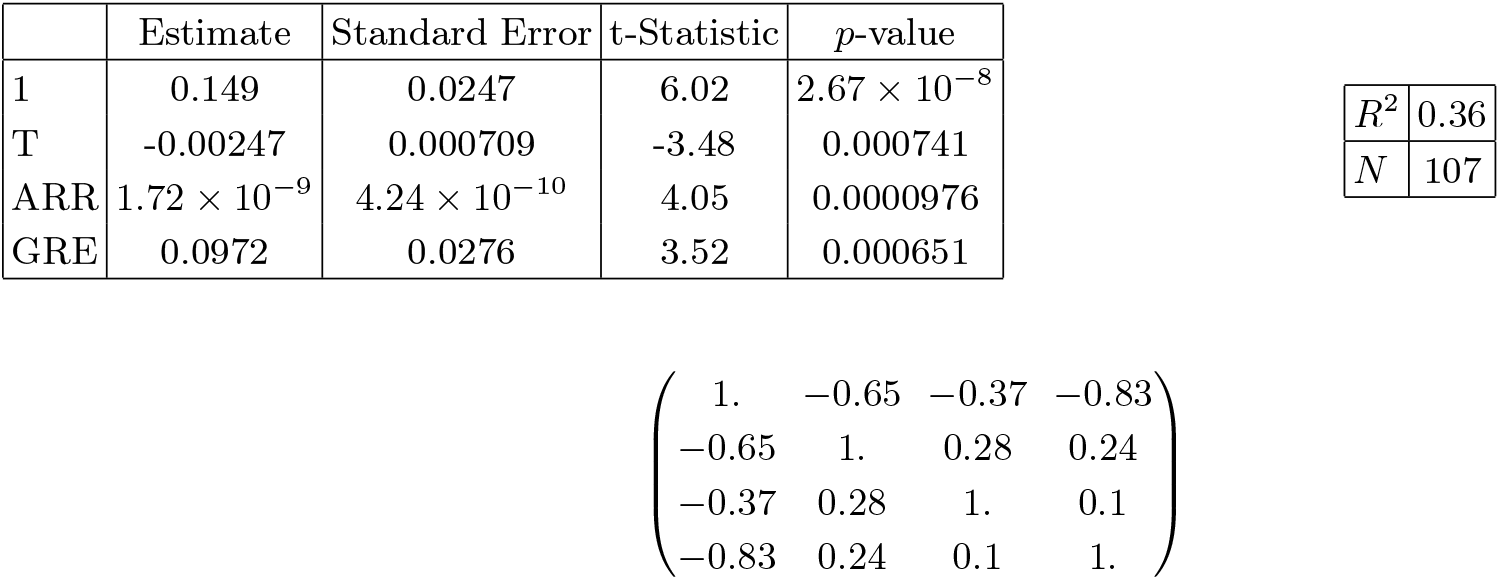
In the left upper panel: best-estimate, standard error (*σ*), t-statistic and *p*-value for the parameters of the linear fit. In the right panels: *R*^2^ for the best-estimate and number of countries *N*. Below we show the correlation matrix for all variables.

| | Estimate | Standard Error | t-Statistic | $p$-value |
| --- | --- | --- | --- | --- |
| 1 | 0.149 | 0.0247 | 6.02 | $2.67 \times 10^{-8}$ |
| T | -0.00247 | 0.000709 | -3.48 | 0.000741 |
| ARR | $1.72 \times 10^{-9}$ | $4.24 \times 10^{-10}$ | 4.05 | 0.0000976 |
| GRE | 0.0972 | 0.0276 | 3.52 | 0.000651 |

|  |  |
| --- | --- |
| $R^2$ | 0.36 |
| $N$ | 107 |

#### B. Blood types

Blood types are not equally distributed in the world and thus we have correlated them with *α*. Data were taken from [50]. Very interestingly we find significant correlations, especially for blood types B+ (slower COVID-19 contagion) and A-(faster COVID-19 contagion). In general also all RH-negative blood types correlate with faster COVID-19 contagion. It is interesting to compare with findings in clinical data: (i) our finding that blood type A is associated with a higher risk for acquiring COVID-19 is in good agreement with [26], (ii) we find higher risk for group 0-and no correlation for group 0+ (while [26] finds lower risk for groups 0), (iii) we have a strong significance for lower risk for RH+ types and in particular lower risk for group B+, which is probably a new finding, to our knowledge. These are also non-trivial findings which should stimulate further medical research on the immune response of different blood-types against COVID-19.

*1. Type A+*

*2. Type B+*

*3. Type 0-*

*4. Type A-*

*5. Type B-*

*6. Type AB-*

*7. RH-positive*

## V. CROSS-CORRELATIONS

In this section we first perform linear fits of *α* with each possible pair of variables (for blood types, we considered only RH+ and B+). We show the correlation coefficients between the two variables, for each pair, in Fig. 30. We also show the *p*-value of the *t*-statistic of each variable in a pair and the total *R*^2^ of such fits in Fig. 31.

**Figure 23:**
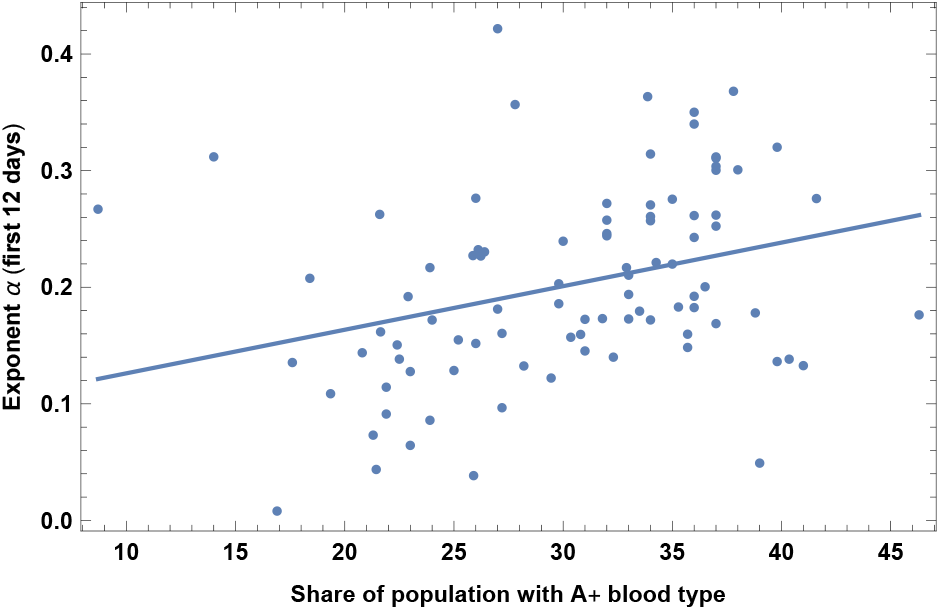
Exponent *α* for each country vs. percentage of population with blood type A+. We show the data points and the best-fit for the linear interpolation.

**Figure 24:**
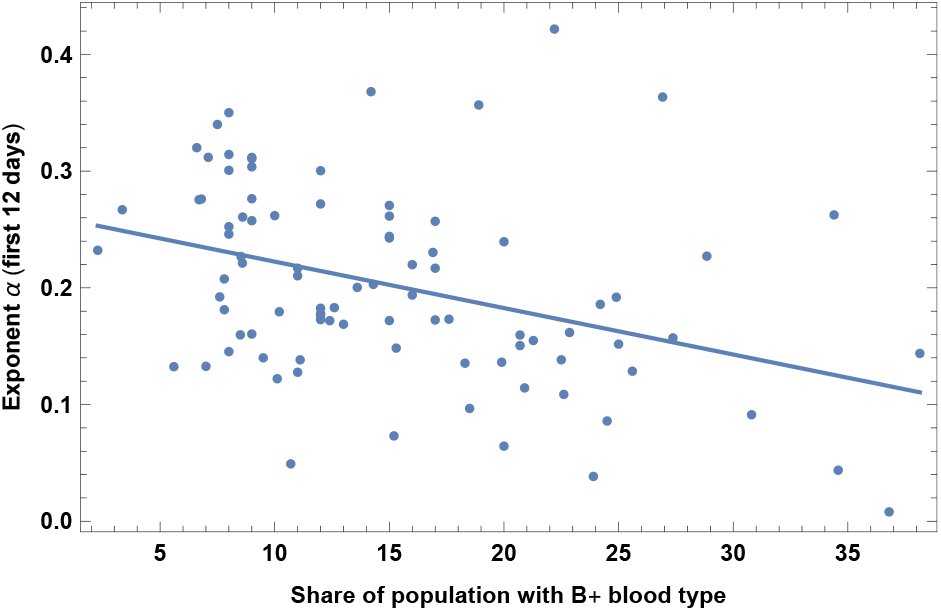
Exponent *α* for each country vs. percentage of population with blood type B+. We show the data points and the best-fit for the linear interpolation.

**Figure 25:**
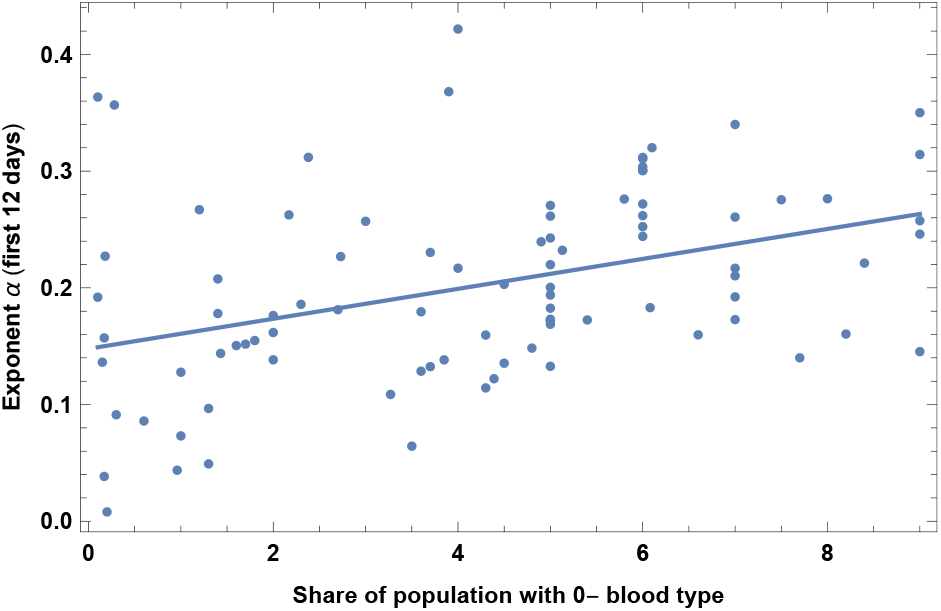
Exponent *α* for each country vs. percentage of population with blood type 0-. We show the data points and the best-fit for the linear interpolation.

**Figure 26:**
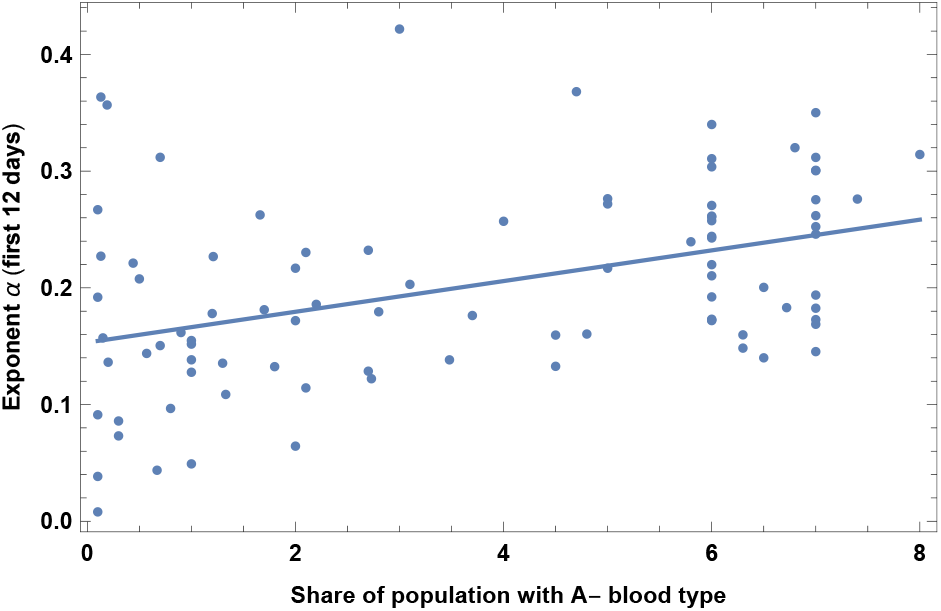
Exponent *α* for each country vs. percentage of population with blood type A-. We show the data points and the best-fit for the linear interpolation.

**Figure 27:**
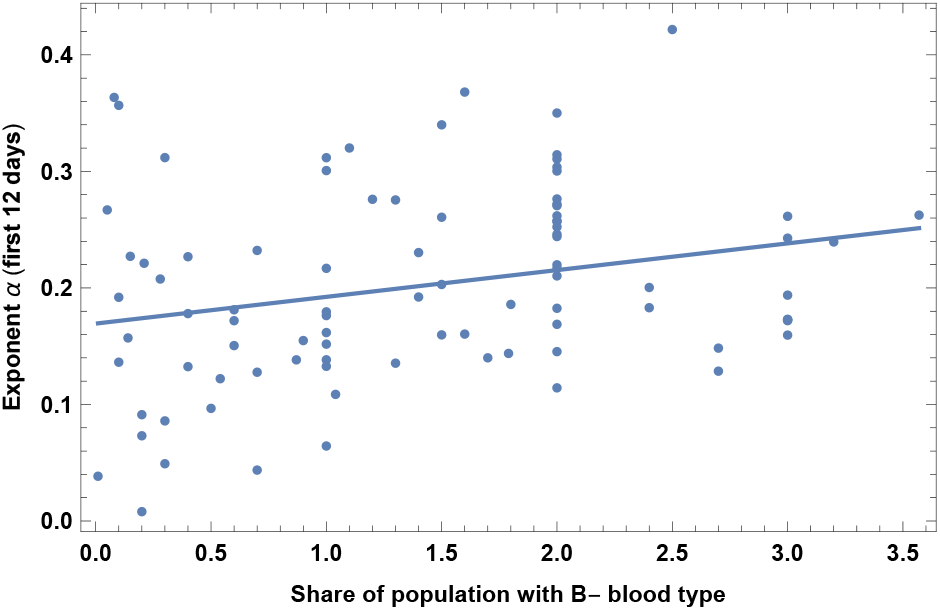
Exponent *α* for each country vs. percentage of population with blood type B-. We show the data points and the best-fit for the linear interpolation.

**Figure 28:**
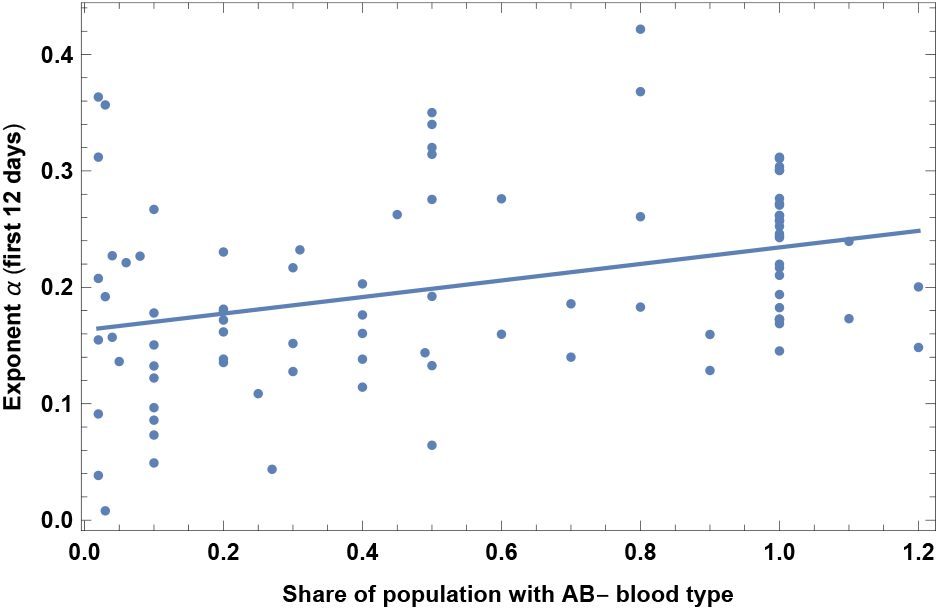
Exponent *α* for each country vs. percentage of population with blood type AB-. We show the data points and the best-fit for the linear interpolation.

**Figure 29:**
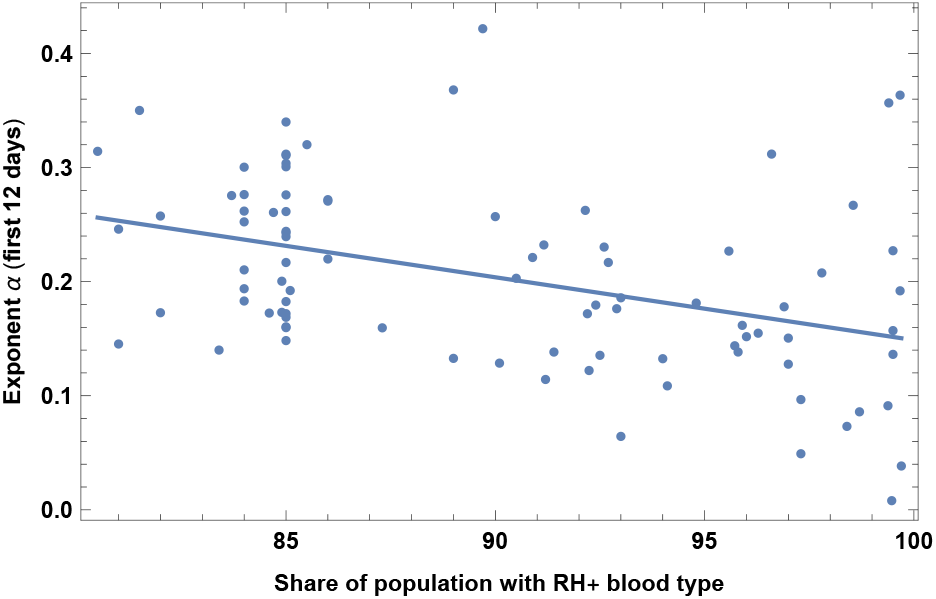
Exponent *α* for each country vs. percentage of population with RH-positive blood. We show the data points and the best-fit for the linear interpolation.

**Figure 30:**
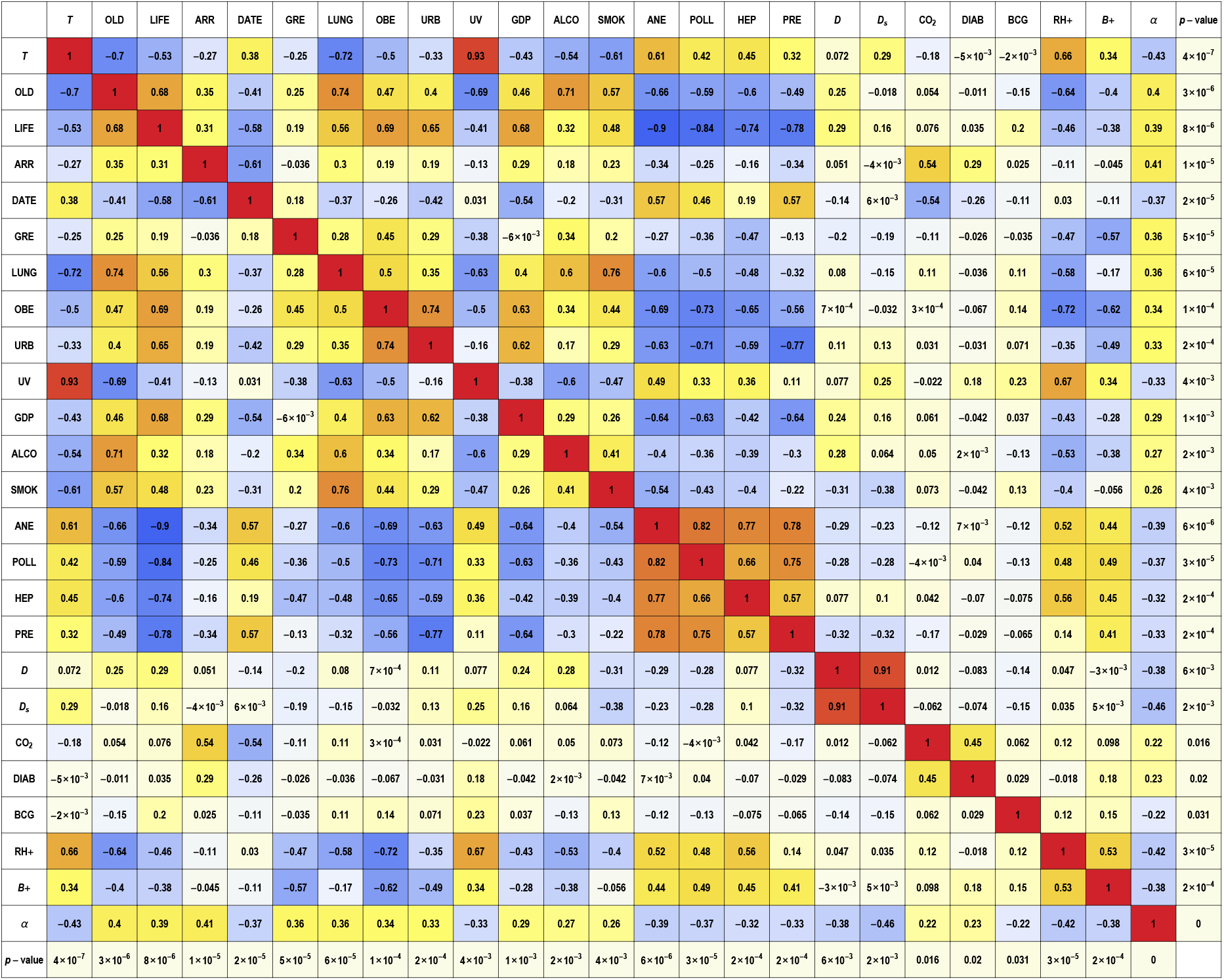
Correlation coefficients between each variable in a pair. Such coefficient corresponds to the off-diagonal entry of the (normalized) covariance matrix, multiplied by −1. In the last column and row we show the *p*-value of each variable when performing a one-variable linear fit for the growth rate *α*. Note also that the fits that include vitamin D variables (D and D_*s*_) and UV index are based on smaller samples than for the other fits and were collected with rather with inhomogeneous data, and so have to be confirmed on a larger sample, as explained in the text. The variables considered here are: Temperature (T), Old age dependency ratio (OLD), Life expectancy (LIFE), Number of tourist arrivals (ARR), Starting date of the epidemic (DATE), Amount of contact in greeting habits (GRE), Lung cancer (LUNG), Obesity in males (OBE), Urbanization (URB), UV Index (UV), GDP per capita (GDP), Alcohol consumption (ALCO), Daily smoking prevalence (SMOK), Prevalence of anemia in children (ANE), Death rate due to pollution (POLL), Prevalence of hepatitis B (HEP), High blood pressure in females (PRE), average vitamin D serum levels (D), seasonal vitamin D serum levels (D_*s*_), CO_2_ emissions (CO_2_), type 1 diabetes prevalence (DIAB), BCG vaccination (BCG), percentage with blood of RH+ type (RH+), percentage with blood type B+ (B+).

**Figure 31:**
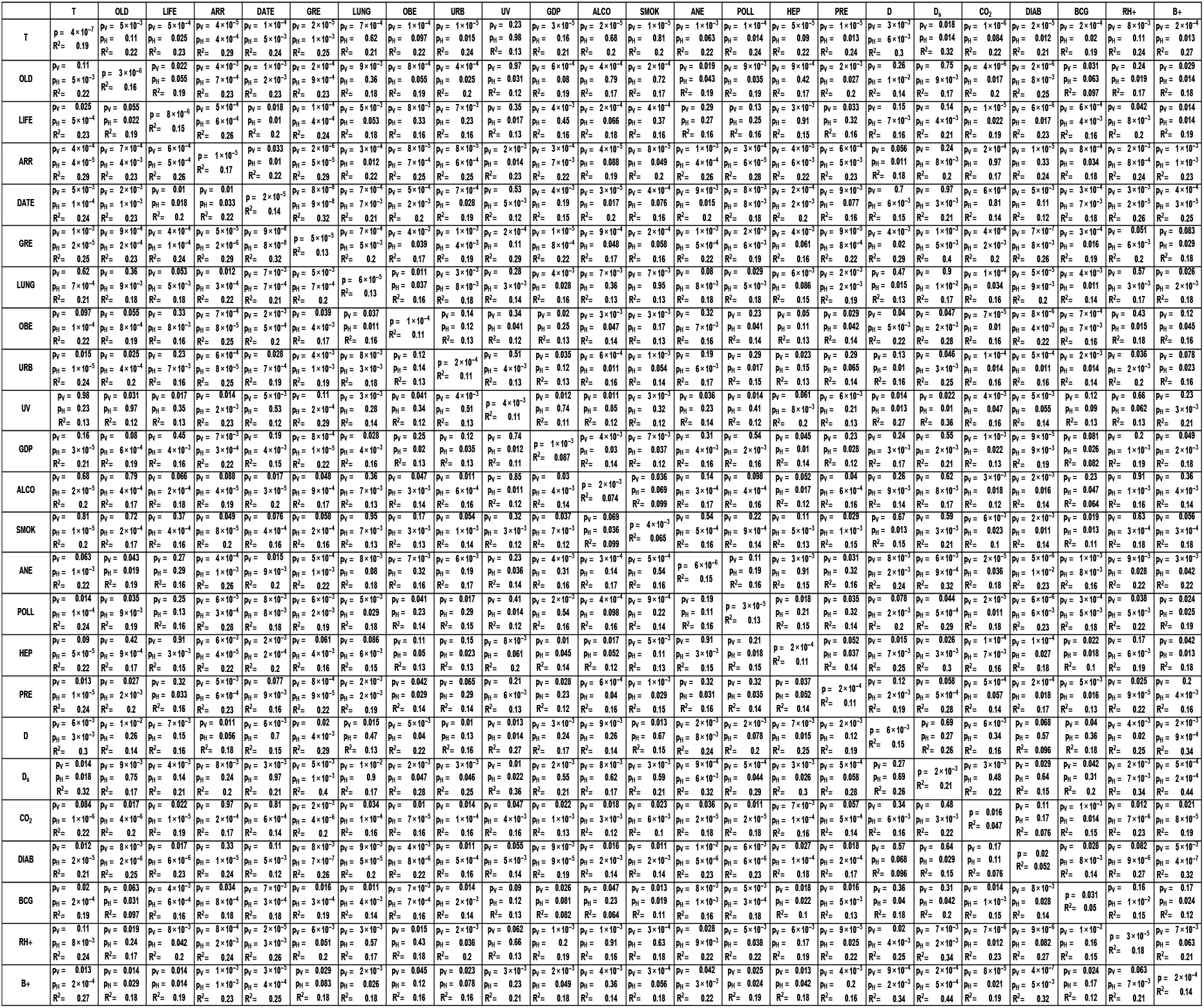
Significance and *R*^2^ for linear fits of *α* as a function of two-variables. Variables names are the same as in Fig. 30. Each cell in the table gives *R*^2^ and the *p*-value of the each of the two variables, using *t*-statistic, labeled as p_*H*_ for horizontal and p_*V*_ for vertical.

We give here below possible interpretations of the redundancy among our variables and we perform multiple variable fits in the following subsections.

### A. Possible Interpretations

The set of most significant variables, i.e. with smallest *p*-value, which correlate with faster propagation of COVID-19 are the following: low temperature, high percentage of old vs. working people and life expectancy, number of international tourist arrivals, high percentage of RH-blood types, earlier starting date of the epidemic, high physical contact in greeting habits, prevalence of lung cancer. Such variables are however correlated with each other and we analyze them together below. Most other variables are mildly/strongly correlated with the previous set of variables, except for: prevalence of Type-I diabetes, BCG vaccination, Vitamin D levels (note however that the latter is based on more inhomogeneous data and should be confirmed on a larger sample), which might indeed be considered as almost independent factors.

From the above table one can verify that all the “counterintuitive” variables (death-rate due to pollution, POLL, prevalence of anemia, ANE, prevalence of hepatitis B, HEP, high blood pressure in women, PRE) have a strong negative correlation with life expectancy, LIFE. This offers a neat possible interpretation: since countries with high deaths by pollution or high prevalence of anemia or hepatitis B or high blood pressure in women have a younger population, then the virus spread is slower. Therefore one expects that when performing a fit with any of such variable and LIFE together, one of the variables will turn out to be non-significant, i.e. redundant. Indeed one may verify from Table 31 that this happens for all of the four above variables.

Redundancy is also present when one of the following variables is used together with life expectancy in a 2 variables fit: smoking, urbanization, obesity in males. Also, old age dependency and life expectancy are obviously quite highly correlated.

Other variables instead do *not* have such an interpretation: BCG vaccination, type-1 diabetes in children and vitamin D levels. In this case other interpretations have to be looked for. Regarding the vaccination a promising interpretation is indeed that BCG-vaccinated people could be more protected against COVID-19 [29–31].

Lung cancer and alcohol consumption remain rather significant, close to a *p*-value of 0.05, even after taking into account life expectancy, but they become non-significant when combining with old age dependency. In this case it is also difficult to disentangle them from the fact that old people are more subject to COVID-19 infection.

Blood type RH+ is also quite correlated with T, however it remains moderately significant when combined with it.

Finally vitamin D, which is measured on a smaller sample, also has little correlations with the main factors. This is quite interesting, since it may open avenues for research on protective factors and health policies. It is quite possible that high Vitamin D helps the immune response to COVID-19 [45–47]. Note however that this finding is based on more inhomogeneous data and should be confirmed on a larger and more homogeneous sample.

### B. Multiple variable fits

It is not too difficult to identify redundant variables, looking at very strongly correlated pairs in Table 30. It is generically harder, instead, to extract useful information when combining more than 2 or 3 variables, since we have many variables with comparable predictive power (individual *R*^2^ are at most around 0.2) and several of them exhibit mild/strong correlations. In the following we perform examples of fits with some of the most predictive variables, trying to keep small correlation between them.

**Table XXXI:** In the left upper panel: best-estimate, standard error (*σ*), t-statistic and *p*-value for the parameters of the linear interpolation. In the right panels: *R*^2^ for the best-estimate and number of countries *N*. Below we show the correlation matrix for all variables.

|  | Estimate | Standard Error | t-Statistic | p-value |  |
| --- | --- | --- | --- | --- | --- |
| 1 | 0.299 | 0.0519 | 5.77 | $8.66 \times 10^{-8}$ | |
| T | -0.00169 | 0.000718 | -2.36 | 0.0203 | $R^2$ 0.42 |
| ARR | $7.69 \times 10^{-10}$ | $4.99 \times 10^{-10}$ | 1.54 | 0.126 | $N$ 107 |
| GRE | 0.13 | 0.0283 | 4.59 | 0.0000127 |  |
| DATE | -0.00237 | 0.000728 | -3.26 | 0.00154 |  |

**Table XXXII:**
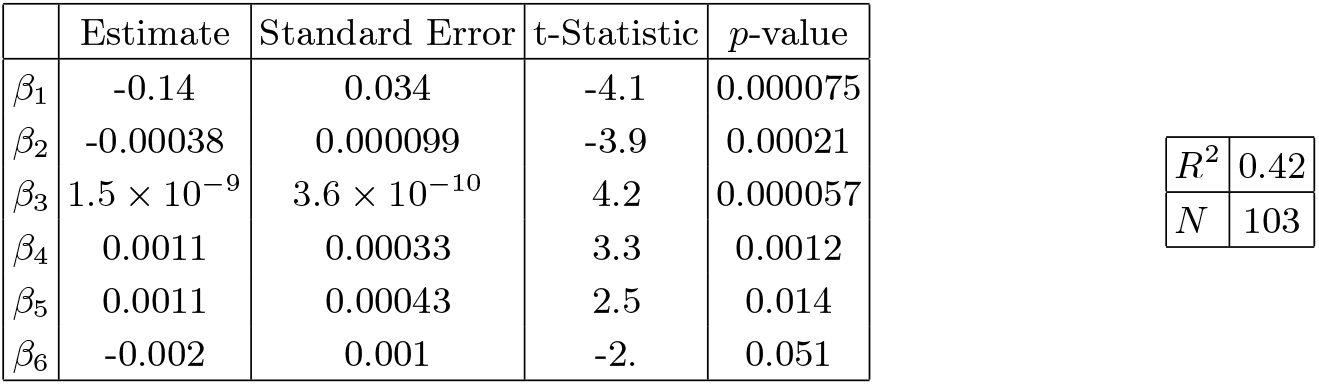
Best-estimate, standard error, t-statistic and *p*-value for the parameters of the Principal Components, see eqs. (1-2). In the right panel: *R*^2^ for the best-estimate and number of countries *N*.

| | Estimate | Standard Error | t-Statistic | $p$-value |
| --- | --- | --- | --- | --- |
| $\beta_1$ | -0.14 | 0.034 | -4.1 | 0.000075 |
| $\beta_2$ | -0.00038 | 0.000099 | -3.9 | 0.00021 |
| $\beta_3$ | $1.5 \times 10^{-9}$ | $3.6 \times 10^{-10}$ | 4.2 | 0.000057 |
| $\beta_4$ | 0.0011 | 0.00033 | 3.3 | 0.0012 |
| $\beta_5$ | 0.0011 | 0.00043 | 2.5 | 0.014 |
| $\beta_6$ | -0.002 | 0.001 | -2. | 0.051 |

#### 1. Temperature+Arrivals+Greetings

Here we show an example of a fit with 3 parameters.

#### B. Temperature+Arrivals+Greetings+Starting date

Here we show an example of a fit with 4 parameters. As we combine more than 3 parameters, typically at least one of them becomes less significant.

#### 3. Many variables fit

One may think of combining *all* our variables. This is however not a straightforward task, because we do not have data on the same number *N* of countries for all variables. As a compromise we may restrict to a large number of variables, but still keeping a large number of countries. For instance we may choose the following set: T, OLD, LIFE, ARR, DATE, GRE, LUNG, OBE, URB, GDP, ALCO, SMOK, ANE, POLL, HEP, PRE, CO_2_. These variables are defined for a sample of *N* = 103 countries. By combining all of them we get *R*^2^ = 0.48, which tells us that only about half of the variance is described by these variables. However, clearly, many of these variables are redundant. We perform thus now a Principal Component Analysis (PCA), i.e. we look for linear combinations of such variables that diagonalize the covariance matrix.

We perform the PCA analysis in two different ways:

1. First, we we exclude the variables with a counterintuitive behavior, see section IV A. Moreover we also exclude LIFE, since its meaning is also captured by the similar variable OLD. We performed thus a Principal Component Analysis, fitting with linear combinations of: 1, T, OLD, ARR, GRE, OBE, URB, SMOK, CO_2_, DATE, LUNG, GDP, ALCO.
2. Second, we include also the “counterintuitive” variables, using thus linear combinations of: 1, T, OLD, ARR, GRE, OBE, URB, SMOK, CO_2_, DATE, LUNG, GDP, ALCO, ANE, HEP, POLL, PRE;

In both cases we fit with

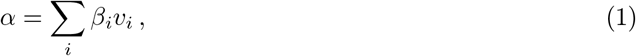

where *v*_*i*_ are linear combinations of the above variables.

In case (1) we find a fit with total *R*^2^ = 0.46, with *N* = 103. There are however only 6 significant independent orthogonal linear combinations. Such 6 combinations, which have *R*^2^ = 0.42, are:

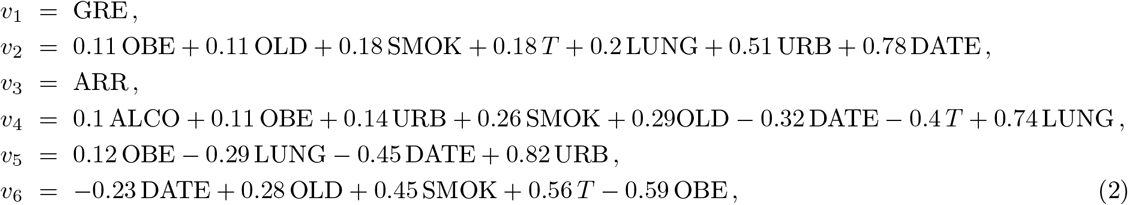

where we omitted variables whose coefficients (“loadings”) on *v*_1_, *v*_2_, *v*_3_, *v*_4_, *v*_6_ are less than 0.1. The variables with loadings of at least 0.3 are: GRE, URB, DATE, ARR, T, LUNG, SMOK, OBE. The significance of the principal components is given in Table XXXII.

In case (2) we find a fit with total *R*^2^ = 0.48, with *N* = 103. There are again only 6 significant independent orthogonal linear combinations. Such 6 combinations, which have *R*^2^ = 0.42, are:

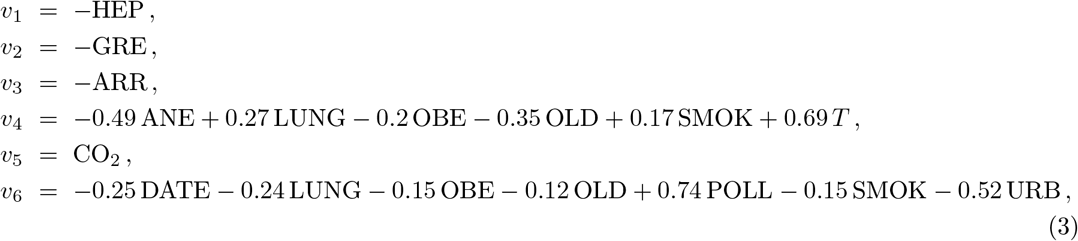

where, again, we omitted variables whose loadings are less than 0.1. The variables with loadings of at least 0.3 are now: HEP, GRE, ARR, ANE, OLD, T, CO_2_, URB. The significance of the principal components is given in Table XXXIII.

**Table XXXIII:**
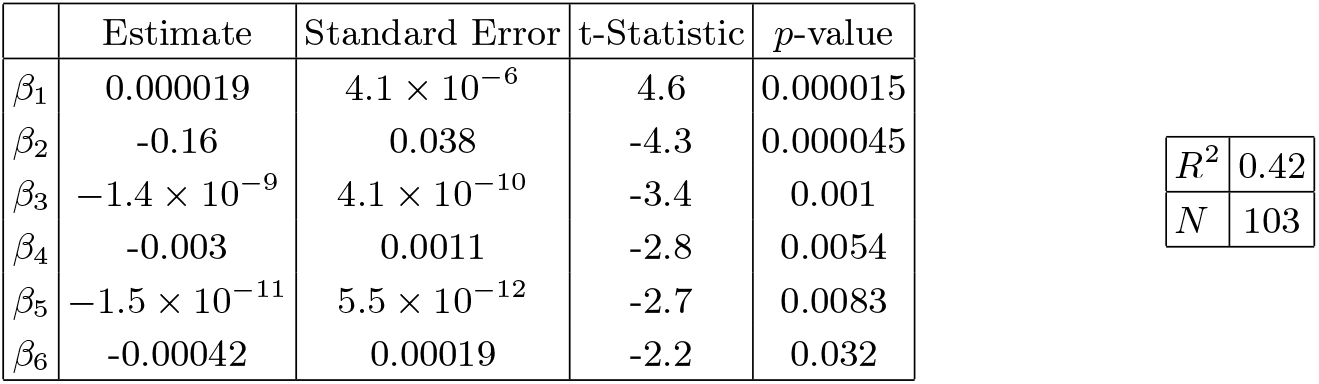
Best-estimate, standard error, t-statistic and *p*-value for the parameters of the Principal Components, see eqs. (1-3). In the right panel: *R*^2^ for the best-estimate and number of countries *N*.

| | Estimate | Standard Error | t-Statistic | $p$-value |
| --- | --- | --- | --- | --- |
| $\beta_1$ | 0.000019 | $4.1 \times 10^{-6}$ | 4.6 | 0.000015 |
| $\beta_2$ | -0.16 | 0.038 | -4.3 | 0.000045 |
| $\beta_3$ | $-1.4 \times 10^{-9}$ | $4.1 \times 10^{-10}$ | -3.4 | 0.001 |
| $\beta_4$ | -0.003 | 0.0011 | -2.8 | 0.0054 |
| $\beta_5$ | $-1.5 \times 10^{-11}$ | $5.5 \times 10^{-12}$ | -2.7 | 0.0083 |
| $\beta_6$ | -0.00042 | 0.00019 | -2.2 | 0.032 |

## VI. CONCLUSIONS

We have collected data for countries that had at least 12 days of data after a starting point, which we fixed to be at the threshold of 30 confirmed cases. We considered a dataset of 126 countries, collected on April 15th. We have fit the data for each country with an exponential and extracted the exponents *α*, for each country. Then we have correlated such exponents with several variables, one by one.

We found a positive correlation with *high confidence* level with the following variables, with respective *p*-value: low temperature (negative correlation, *p*-value 4 ·10^−7^), high ratio of old people vs. people in the working-age (15-64 years) (*p*-value 3· 10^−6^), life expectancy (*p*-value 8· 10^−6^), international tourism: number of arrivals (*p*-value 1·10^−5^), earlier start of the epidemic (*p*-value 2· 10^−5^), high amount of contact in greeting habits (positive correlation, *p*-value 5 ·10^−5^), lung cancer death rates (*p*-value 6· 10^−5^), obesity in males (*p*-value 1 ·10^−4^), share of population in urban areas (*p*-value 2 · 10^−4^), share of population with cancer (*p*-value 2.8·10^−4^), alcohol consumption (*p*-value 0.0019), daily smoking prevalence (*p*-value 0.0036), low UV index (*p*-value 0.004; smaller sample, 73 countries), low vitamin D serum levels (annual values *p*-value 0.006, seasonal values 0.002; smaller sample, ∼50 countries).

We also find strong evidence for correlation with blood types: RH + blood group system (negative correlation, *p*-value 3 ·10^−5^); A+ (positive correlation, *p*-value 3 · 10^−3^); B + (negative correlation, *p*-value 2 ·10^−4^); A-(positive correlation, *p*-value 3· 10^−5^); 0-(positive correlation, *p*-value 8· 10^−4^); AB-(positive correlation, *p*-value 0.028); We find moderate evidence for correlation with: B-(positive correlation, *p*-value 0.013).

We find *moderate* evidence for positive correlation with: CO_2_ (and SO) emissions (*p*-value 0.015), type-1 diabetes in children (*p*-value 0.023), vaccination coverage for Tuberculosis (BCG) (*p*-value 0.028).

Counterintuitively we also find negative correlations, in a direction opposite to a naive expectation, with: death rate from air pollution (*p*-value 3 ·10^−5^), prevalence of anemia, adults and children, (*p*-value 1 ·10^−4^ and 7 ·10^−6^, respectively), share of women with high-blood-pressure (*p*-value 2 ·10^−4^), incidence of Hepatitis B (*p*-value 2 ·10^−4^), PM2.5 air pollution (*p*-value 0.029).

As is clear from the figures, the data present a high amount of dispersion, for all fits that we have performed. This is of course unavoidable, given the existence of many systematic effects. One obvious factor is that the data are collected at *country* level, whereas many of the factors considered are regional. This is obvious from empirical data (see for instance the difference between the epidemic development in Lombardy vs. other regions in Italy, or New York vs. more rural regions), and also sometimes has obvious explanations (climate, health factors vary a lot region by region) as well as not so obvious ones. Because of this, we consider *R*^2^ values as at least as important as *p*-values and correlation coefficients: an increase of the *R*^2^ after a parameter is included means that the parameter has a systematic effect in reducing the dispersion (“more data points are explained”).

Several of the above variables are correlated with each other and so they are likely to have a common interpretation and it is not easy to disentangle them. The correlation structure is quite rich and non-trivial, and we encourage interested readers to study the tables in detail, giving both *R*^2^, *p*-values and correlation estimates. Note that some correlations are “obvious”, for example between temperature and UV radiation. Others are accidental, historical and sociological. For instance, social habits like alcohol consumption and smoking are correlated with climatic variables. In a similar vein correlation of smoking and lung cancer is very high, and this is likely to contribute to the correlation of the latter with climate. Historical reasons also correlate climate with GDP per capita.

Other variables are found to have a counterintuitive *negative* correlation, which can be explained due their strong negative correlation with life expectancy: death-rate due to pollution, prevalence of anemia, Hepatitis B and high blood pressure for women.

We also analyzed the possible existence of a bias: countries with low GDP-per capita, typically located in warm regions, might have less intense testing and we discussed the correlation with the above variables, showing that most of them remain significant, even after taking GDP into account. In this respect, note that in countries where testing is not prevalent, registration of the illness is dependent on the development of severe symptoms. Hence, while this study is about *infection rates* rather than *mortality*, in quite a few countries we are actually measuring a proxy of mortality rather than infection rate. Hence, effects affecting mortality will be more relevant. Pre-existing lung conditions, diabetes, smoking and health indicators in general as well as pollution are likely to be important in this respect, perhaps not affecting *α* per se but the detected amount of *α*. These are in turn generally correlated with GDP and temperature for historical reasons. Other interpretations, which may be complementary, are that co-morbidities and old age affect immune response and thus may directly increase the growth rate of the contagion. Similarly it is likely that individuals with co-morbidities and old age, developing a more severe form of the disease, are also more contagious than younger or asymptomatic individuals, producing thus an increase in *α*. In this regard, we wish to point the reader’s attention to the relevant differences in correlations once we apply a threshold on GDP per capita. It has long been known that human wellness (we refer to a psychological happiness study [54], but the point is more general) depends non-linearly on material resources, being strongly correlated when resources are low and reaching a plateau after a critical limit. The biases described above (weather comorbidity, testing facilities, pre-existing conditions and environmental factors) seem to reflect this, changing considerably in the case our sample has a threshold w.r.t. a more general analysis without a threshold.

About pollution our findings are mixed. We find no correlation with generic air pollution (“Suspended particulate matter (SPM), in micrograms per cubic metre”). We find higher contagion to be moderately correlated only with and CO_2_/SO emissions. Instead we find a *negative* correlation with death rates due to air pollution and PM2.5 concentration (in contrast with [48], see also [55]). Note however that correlation with PM2.5 becomes non significant when combined with GDP per capita, while CO_2_/SO becomes non significant when combined with tourist arrivals. Finally death rates due to air pollution is also redundant when correlating with life expectancy.

We also performed a Principal Component Analysis, in a sample of N=103 countries, where: (1) we omitted all variables with a counterintuitive behavior, (2) we omitted Vitamin D, BCG, UV and Blood variables, as we did not have data for so many countries, (3) we also omitted LIFE, since its meaning is already described by the similar variable OLD. Therefore we used the following variables: 1, T, OLD, ARR, GRE, OBE, URB, SMOK, CO2, DATE, LUNG, GDP, ALCO. As a result we found that only 6 linear orthogonal combinations are significant, and the variables with a loading of at least 0.3 on any of such 6 combinations are: GRE, URB, DATE, ARR, T, LUNG, SMOK, OBE. Including also the counterintuitive variables we find again only 6 significant linear orthogonal combinations, but the variables with a loading of at least 0.3 are now: HEP, GRE, ARR, ANE, OLD, T, CO_2_, URB.

Some of the variables that we have studied cannot be arbitrarily changed, but can be taken into account by public health policies, such as temperature, amount of old people and life expectancy, by implementing stronger testing and tracking policies, and possibly lockdowns, both with the arrival of the cold seasons and for the old aged population.

Other variables instead can be controlled by governments: testing and isolating international travelers and reducing number of flights in more affected regions; promoting social distancing habits as long as the virus is spreading, such as campaigns for reducing physical contact in greeting habits; campaigns against vitamin D deficiency, decrease smoking and obesity.

We also emphasize that some variables are useful to inspire and support medical research, such as correlation of contagion with: lung cancer, obesity, low vitamin D levels, blood types (higher risk for all RH-types, A types, lower risk for B+ type), type 1 diabetes. This definitely deserves further study, also of correlational type using data from patients.

In conclusion, our findings could thus be very useful both for policy makers and for further experimental research.

## Data Availability

All data are publicly available

## Acknowledgments

GT acknowledges support from FAPESP proc. 2017/06508-7, partecipation in FAPESP tematico 2017/05685-2 and CNPQ bolsa de produtividade 301432/2017-1. We would like to acknowledge Alberto Belloni, Jordi Miralda and Miguel Quartin for useful discussions and comments.

## Appendix A: Vitamin D

We collected most data on vitamin D from [35–39] and from references therein. For a first dataset of 50 countries we have collected annual averages. For many countries several studies with different values were found and in this case we have collected the mean and the standard error (when available) and a weighted average has been performed. The resulting values that we have used are listed in Table XXXIV. The sample size here is smaller and such dataset is based on quite inhomogeneous research and thus should be confirmed by a more complete dataset.

**Table XXXIV:**
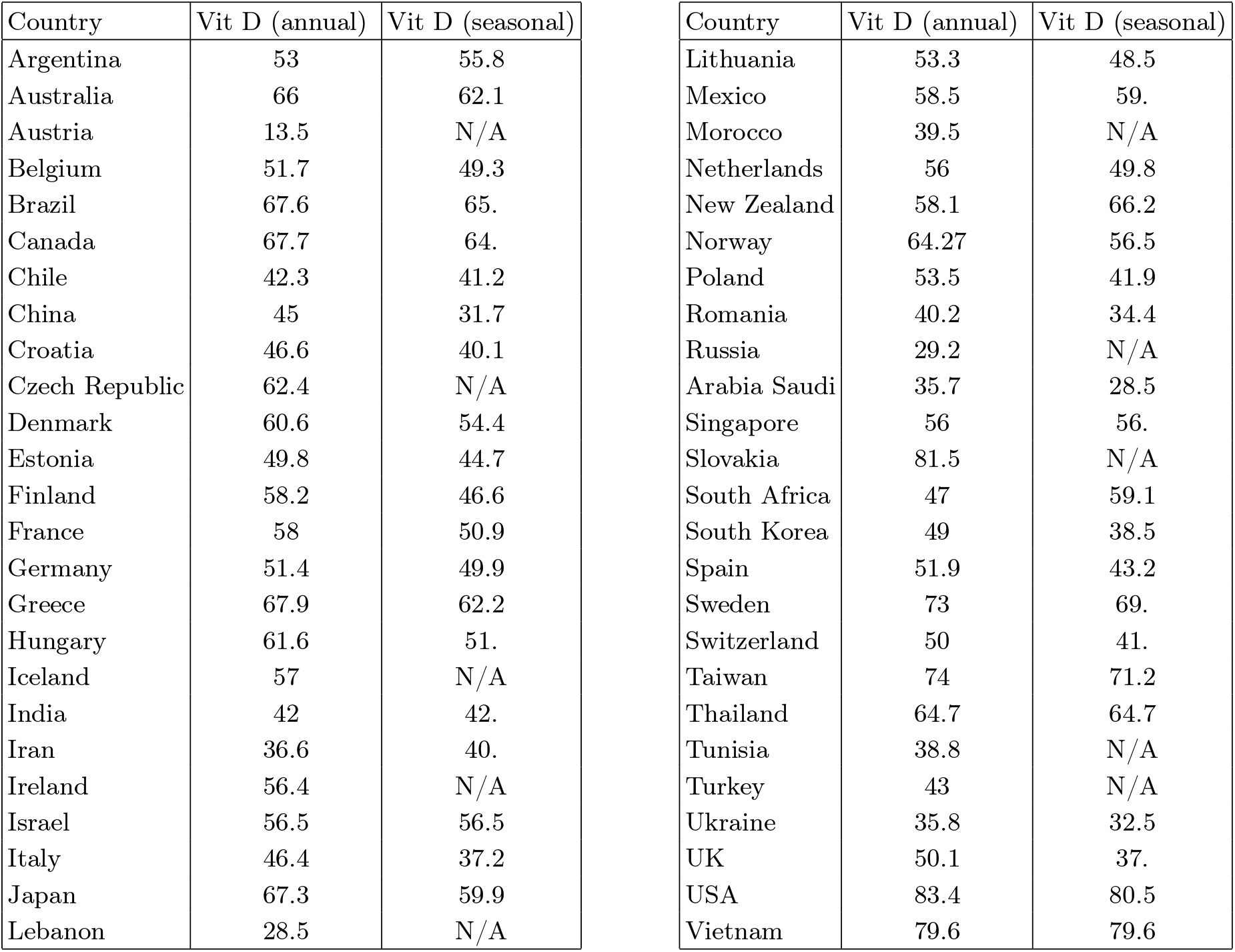
Vitamin D serum levels (in nmol/l) obtained with a weighted average from refs. [35–39] and references therein. The “annual” level refers to an average over the year. The “seasonal” level refers to the value present in the literature, which is closer to the months of January-March: either the amount during such months or during winter for northern hemisphere, *or* during summer for southern hemisphere *or* the annual level for countries with little seasonal variation.

| Country | Vit D (annual) | Vit D (seasonal) | Country | Vit D (annual) | Vit D (seasonal) |
| --- | --- | --- | --- | --- | --- |
| Argentina | 53 | 55.8 | Lithuania | 53.3 | 48.5 |
| Australia | 66 | 62.1 | Mexico | 58.5 | 59. |
| Austria | 13.5 | N/A | Morocco | 39.5 | N/A |
| Belgium | 51.7 | 49.3 | Netherlands | 56 | 49.8 |
| Brazil | 67.6 | 65. | New Zealand | 58.1 | 66.2 |
| Canada | 67.7 | 64. | Norway | 64.27 | 56.5 |
| Chile | 42.3 | 41.2 | Poland | 53.5 | 41.9 |
| China | 45 | 31.7 | Romania | 40.2 | 34.4 |
| Croatia | 46.6 | 40.1 | Russia | 29.2 | N/A |
| Czech Republic | 62.4 | N/A | Arabia Saudi | 35.7 | 28.5 |
| Denmark | 60.6 | 54.4 | Singapore | 56 | 56. |
| Estonia | 49.8 | 44.7 | Slovakia | 81.5 | N/A |
| Finland | 58.2 | 46.6 | South Africa | 47 | 59.1 |
| France | 58 | 50.9 | South Korea | 49 | 38.5 |
| Germany | 51.4 | 49.9 | Spain | 51.9 | 43.2 |
| Greece | 67.9 | 62.2 | Sweden | 73 | 69. |
| Hungary | 61.6 | 51. | Switzerland | 50 | 41. |
| Iceland | 57 | N/A | Taiwan | 74 | 71.2 |
| India | 42 | 42. | Thailand | 64.7 | 64.7 |
| Iran | 36.6 | 40. | Tunisia | 38.8 | N/A |
| Ireland | 56.4 | N/A | Turkey | 43 | N/A |
| Israel | 56.5 | 56.5 | Ukraine | 35.8 | 32.5 |
| Italy | 46.4 | 37.2 | UK | 50.1 | 37. |
| Japan | 67.3 | 59.9 | USA | 83.4 | 80.5 |
| Lebanon | 28.5 | N/A | Vietnam | 79.6 | 79.6 |

